# Personalized Single-cell Transcriptomics Reveals Molecular Diversity in Alzheimer’s Disease

**DOI:** 10.1101/2024.11.01.24316589

**Authors:** Pramod Bharadwaj Chandrashekar, Sayali Anil Alatkar, Noah Cohen Kalafut, Ting Jin, Chirag Gupta, Ryan Burzak, Xiang Huang, Shuang Liu, Athan Z. Li, PsychAD Consortium, Kiran Girdhar, Georgios Voloudakis, Gabriel E. Hoffman, Jaroslav Bendl, John F. Fullard, Donghoon Lee, Panos Roussos, Daifeng Wang

**Author notes:** Co-first authors (Pramod Bharadwaj Chandrashekar, Sayali Anil Alatkar, Noah Cohen Kalafut, Ting Jin).

## Abstract

Precision medicine for brain diseases faces many challenges, including understanding the heterogeneity of disease phenotypes. Such heterogeneity can be attributed to the variations in cellular and molecular mechanisms across individuals. However, personalized mechanisms remain elusive, especially at the single-cell level. To address this, the PsychAD project generated population-level single-nucleus RNA-seq data for 1,494 human brains with over 6.3 million nuclei covering diverse clinical phenotypes and neuropsychiatric symptoms (NPSs) in Alzheimer’s disease (AD). Leveraging this data, we analyzed personalized single-cell functional genomics involving cell type interactions and gene regulatory networks. In particular, we developed a knowledge-guided graph neural network model to learn latent representations of functional genomics (embeddings) and quantify importance scores of cell types, genes, and their interactions for each individual. Our embeddings improved phenotype classifications and revealed potentially novel subtypes and population trajectories for AD progression, cognitive impairment, and NPSs. Our importance scores prioritized personalized functional genomic information and showed significant differences in regulatory mechanisms at cell type level across various phenotypes. Such information also allowed us to further identify subpopulation-level biological pathways, including ancestry for AD. Finally, we associated genetic variants with cell type-gene regulatory network changes across individuals, i.e., gene regulatory QTLs (grQTLs), providing novel functional genomic insights compared to existing QTLs. We further validated our results using external cohorts. Our analyses are available through iBrainMap, an open-source computational framework, and as a personalized functional genomic atlas for Alzheimer’s Disease.

## Main

Precision medicine in neuropsychiatric and neurodegenerative diseases considers variability in genomics, environment, and lifestyle for each person in order to dissect the complex and heterogeneous etiology of each disorder, and to deliver more accurate diagnoses and targeted therapies^1–4^. The heterogeneity of these diseases stems from differences in cellular and molecular mechanisms. Functional genomics examines cell type interactions and cell type gene regulatory mechanisms to dissect this heterogeneity and to uncover disease mechanisms^2^. To advance precision medicine in AD and to further our understanding of disease heterogeneity, there is an urgent need to perform personalized analysis that better captures interindividual variability. Current personalized analyses in brain diseases predominantly rely on brain imaging^5–8^, often overlooking crucial biologically mechanistic information. Recently, several attempts have been made to capture disease-specific and personalized gene expression patterns^9–11^. For example, Dozier^11^ identified individual variations in immune response among AD using personalized gene co-expression network analysis. Another study identified personalized network patterns and driver genes in various cancers by building individual-specific networks from gene expression data^9^. However, gene expression is fundamentally governed by gene regulation, which is a highly cell-type-specific molecular mechanism. Similarly, several population studies have shown that intercellular crosstalk is an essential component in the process of AD (e.g., astrocyte-microglia crosstalk in β-amyloid pathology^12^ and neuroinflammation^13^). This suggests that capturing personalized cell type interactions and gene regulatory mechanisms at the single-cell level can provide mechanistic insights into disease development, heterogeneity, and progression^9,14,15^.

Large amounts of individual data are essential to capture the complexities and variability present across populations in AD. Recent developments in single-cell sequencing technology have provided opportunities to characterize cell diversity, reveal the genomic landscape, and understand disease heterogeneity of the human brain at single-cell resolution^16–18^. To explore neurodegenerative and neuropsychiatric disease mechanisms, the PsychAD Consortium (**Supplementary Notes “PsychAD dataset”**) generated population-level single-nucleus RNA sequencing (snRNA-seq) data from 1,494 donors across 6.3 million nuclei from the human dorsolateral prefrontal cortex (DLPFC) brain region^19,20^. The dataset includes donors with neurodegenerative or neuropsychiatric disorders such as AD, schizophrenia (SCZ), and diffuse Lewy body disease (DLBD), either as standalone diagnoses or in combinations of multiple diagnoses. For a subset of samples, there is detailed information on neuropsychiatric symptoms (NPS, e.g., depression). Additionally, quantitative measurements of AD-related phenotypes, including Braak stages (neurofibrillary tangles) and cognitive impairment, are available for all donors diagnosed with AD and the majority of neurotypical controls as well.

Analyzing such large-scale population data typically requires emerging computational approaches to discover the personalized functional genomic information that traditional methods cannot capture. Existing machine learning approaches to analyze AD-associated single cell data have focused on identifying key genes and biomarkers, reconstructing cellular trajectories underlying neurodegeneration, and conducting multi-omics integrative analyses to study the disease in detail^16,21,22^. More advanced approaches like graph neural network based methods^23^ pool cells from the population to learn cell embeddings which are then used for cell clustering^24,25^, inferring phenotypic biological networks^24,26^, and multimodal integration^24–26^. These approaches mainly focus on group-level analyses and are, therefore, effective in identifying group level gene expression and cell embedding patterns. However, they often fail to capture individual-specific patterns, potentially overlooking functional genomics that may be crucial for some individuals but are diluted in population-wide averages.

Here, we present a personalized single-cell transcriptomics analysis of 1,494 human brains from the PsychAD cohort. Our analysis reveals personalized functional genomic patterns, prioritizing cell types, genes and their interactions for AD and clinical phenotypes. We demonstrated the robustness of our results using independent cohorts. We also identify genetic variants associated with cell-type-level gene regulation for 27 cell subclasses. Our results are accessible online through a personalized functional genomic atlas and a computational framework for general personalized analyses.

### Personalized functional genomic atlas across brain disease phenotypes

To perform personalized analyses, we developed a computational framework called iBrainMap that leverages snRNA-seq from multiple donors for phenotype classification, population subtyping, and the prioritization of functional genomic information at the individual level (Figure 1a). We applied this framework to data from the PsychAD consortium^19^, which includes over 6.3 million nuclei (including 27 cell types) from 1,494 donor brains, representing a diverse range of cross-disorders, AD progression phenotypes, and a range of AD-associated NPSs. iBrainMap constructs Personalized Functional Genomics graphs (PFGs) for each donor, integrating prior biological knowledge of diseases into each graph. Each PFG is a directed graph, with nodes representing cell types, transcription factor genes (TFs), and target genes (TGs), and edges denoting cell type interactions and cell type regulatory links from TFs to TGs. These PFGs are then used as input for a knowledge-guided graph neural network (KG-GNN), generating two key outputs for each individual: graph embeddings and importance scores for PFG nodes and edges, facilitating several downstream analyses (see **Methods, Supplementary Note 1-2**).

**Figure 1:**
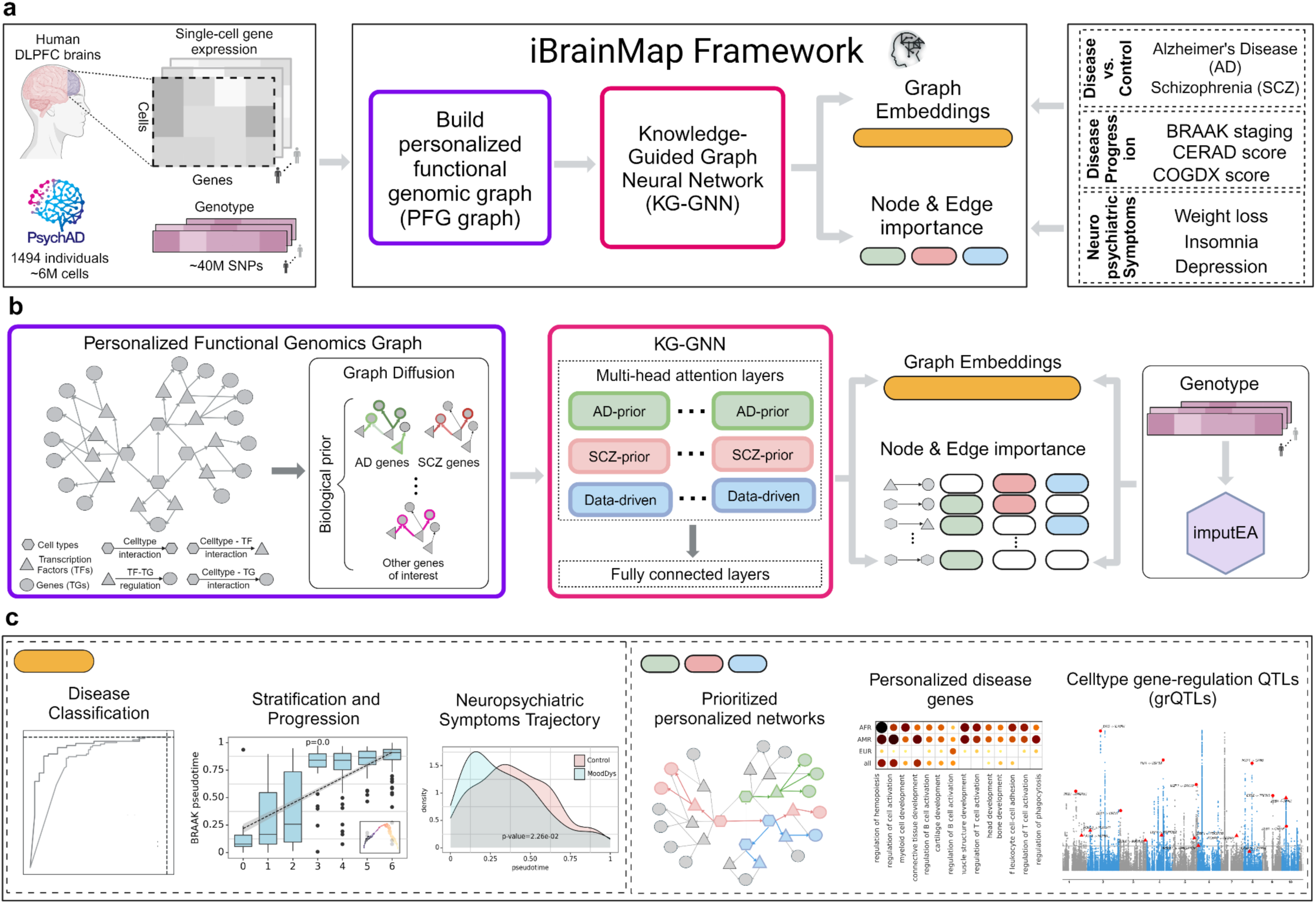
Personalized functional genomic atlas and analysis for brain disease phenotypes. **a,** The iBrainMap framework uses the PsychAD^20^ snRNA-seq data of 1,494 human brain donors to output personalized disease-associated functional genomics information for brain diseases including Alzheimer’s Disease (AD) and Schizophrenia (SCZ). **b**, The iBrainMap framework first constructs a personalized functional genomics graph (PFG) and then applies graph diffusion to integrate known disease genes to generate bio-diffused PFGs for each individual. The KG-GNN, a multi-head graph attention model, uses the bio-diffused PFGs of each individual to classify AD vs. Control, learn graph embeddings, and prioritize nodes and edges for AD (through learned AD, SCZ, and data-driven priors). Genotype data can be used to capture variations in our model and impute donor graph embeddings. These imputed embeddings can then be used in cohorts that only have genetic information for phenotype predictions. More details in **Methods**, **Supplementary Note 1-2, Supplementary** Figures 1-2, **c,** Using graph embeddings, we can classify donors across disease phenotypes and stratify them into potential novel subtypes. Using prioritized edges, we can prioritize phenotype-associated personalized networks, identify known and novel disease genes, and perform cell type-level gene-regulation QTLs (grQTLs) that link associated SNPs with gene regulatory links.

We enhance the PFGs by incorporating prior biological knowledge of known disease genes (e.g., AD and SCZ genes^27,28^), resulting in *bio-diffused* PFGs where edges associated with disease gene nodes are assigned higher weights (see **Methods**, Extended Figure 1a-b). These *bio-diffused* PFGs are then input to our KG-GNN model for classification into disease (case) and control groups (see **Methods**, **Supplementary Note 1**, **Supplementary** Figure 1). The KG-GNN model, a multi-head graph attention network, learns two types of priors (or attention scores): data-driven priors from the input data and disease-specific priors from prior knowledge of disease genes (e.g. AD-prior and SCZ-prior) (see **Supplementary Note 2**, **Supplementary** Figure 2). Following classification, the KG-GNN model outputs graph embeddings and importance scores (for PFG nodes and edges) for each individual through learned priors, which are then used to rank disease-associated functional genomics (nodes: cell types, TFs, TGs; edges: cell type to TF, cell type to TG, and TF-TG) across individuals. Additionally, an imputation module leverages genotype information to impute graph embeddings for new individuals enabling disease diagnosis (Figure 1b).

Based on our framework’s outputs, we conducted downstream analyses in two parts (Figure 1c). First, using graph embeddings, we classified donors according to disease-related phenotypes, including case-control status, pathology capturing different stages of AD, and presence of NPSs.

Additionally, we performed population subtyping to identify novel subgroups capturing different disease stages. With pseudotime analyses of the graph embeddings, we uncovered several population trajectories associated with AD pathology, cognitive status, and NPSs. Second, we used importance scores to prioritize nodes and edges of individuals (through learned AD, SCZ, and data-driven priors), which were then utilized to identify significant subnetworks and to uncover potential biomarkers associated with AD phenotypes, including cell types and cell type regulatory elements (TFs, TGs). Additionally, we associated genetic variants with cell type gene regulatory network changes across individuals, i.e., gene regulatory QTLs (grQTLs), providing novel functional genomic insights linking SNPs to TF-TG regulatory mechanism as opposed to linking SNPs to genes and regulatory elements in traditional QTL analysis.

### Phenotype classification and subtyping using personalized functional genomics

To study the functional genomics associated with AD, we trained our KG-GNN model for binary classification of individual *bio-diffused* PFGs into AD and Control groups. Our KG-GNN model demonstrated superior classification performance compared to several state-of-the-art machine learning models. Specifically, we trained the KG-GNN and other models on 80% of donors from the MSSM cohort with AD diagnosis information: AD (n=438) vs. Control (n=179) (see **Supplementary Note 4**). The KG-GNN model achieved a high area-under-the-curve (AUC) of 0.93 on the 20% held-out donors from MSSM with AD (n= 62) vs. Control (n=30). Further validation on independent data with AD (n=93) vs. Controls (n=68), combined from RADC and SEA-AD^17^ cohorts, yielded a combined AUC of 0.808 (Figure 2a**, Supplementary** Figure 3). Benchmarking against state-of-the-art graph learning models showed that KG-GNN outperformed them, achieving a high AUC score (**Supplementary** Figure 4a). The KG-GNN model demonstrated superior performance over traditional machine learning algorithms that use gene expression alone for population disease classification of AD and Controls (**Supplementary** Figure 5, see **Supplementary Note 5**).

**Figure 2:**
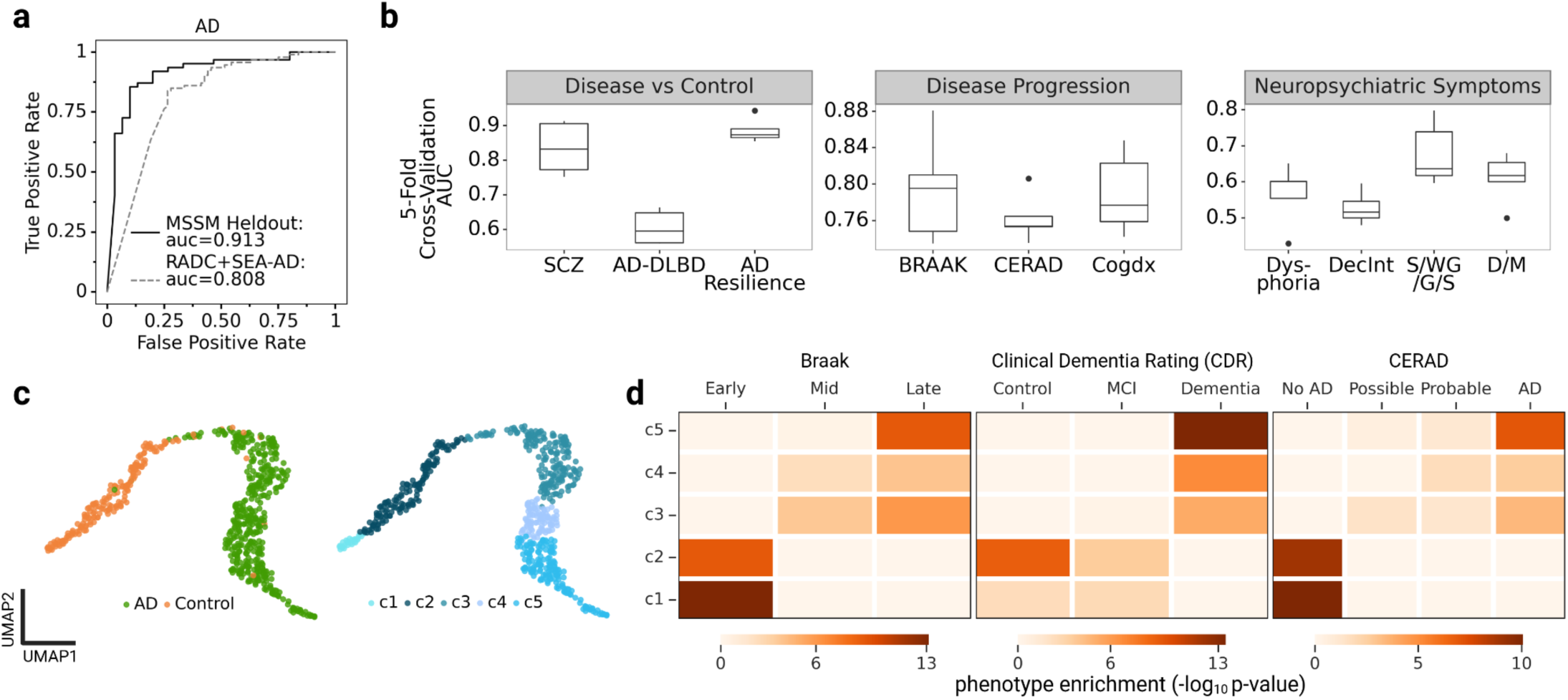
Graph embeddings for personalized functional genomics enable high classification accuracy and disease progression subtyping. **a,** ROC curves for classifying KG-GNN graph embeddings for AD vs. Controls from the MSSM held-out: AD (n=62) vs. Control (n=30) and the independent validation data RADC+SEA-AD: AD (n=93) vs. Controls (n=68). The datasets are described in Extended Figure 1c. **b,** Average five-fold cross-validated ROC for classification of KG-GNN graph embeddings across phenotype contrasts from Extended Figure 1c. **c,** Graph embeddings based UMAP (Left: colored by AD donors vs Controls; Right: colored by clusters from unsupervised clustering/subtyping of graph embeddings) **d,** Heatmaps depict phenotype enrichments for clusters from **(c)** across AD progressions: AD pathology (Braak, CERAD) and cognitive status (clinical dementia rating) showing an increasing trend from c1-c5, using -log10(hypergeometric test p-value) (see **Methods**).

Next, we applied the pre-trained KG-GNN model to *bio-diffused* PFGs from donors across other MSSM contrasts other than AD vs. Control (Extended Figure 1c) to extract graph embeddings. These embeddings were then classified using machine learning models for binary (Disease vs. Control, NPS) and multi-class (Disease Progression) classification tasks. Here we report a high cross-validated AUC score for multiple phenotypes (Figure 2b**; Supplementary Note 6**).

Additionally, we used the donor graph embeddings to classify cross-diseases, including SCZ, on an independent cohort from NIMH-IRP Human Brain Collection Core (HBCC): SCZ (n=50) vs. Control (n=250) (**Supplementary** Figure 6) and achieved an ROC of 0.616. These results highlight the robustness and versatility of our framework for disease and phenotype classifications.

Our model effectively distinguished between AD and Controls, as visualized in the UMAP representation of graph embeddings (Figure 2c: left). We further explored potential novel subtypes within these graph embeddings using unsupervised clustering (see **Methods**), identifying five distinct clusters (c1-c5) (Figure 2c: right). Enrichment analysis across AD progression phenotypes revealed that these clusters capture trends in Braak stages, cognition (Clinical Dementia Rating (CDRscore)), and CERAD score (Figure 2d). Specifically, c1-c2 correspond to (a) early Braak stages (Braak 0,1,2), (b) controls in CDRscore, and (c) ‘No AD’ based on CERAD score; while clusters c4-c5 were associated with (a) later Braak stages (Braak 5,6), (b) Dementia in clinical diagnosis of cognitive status (CDRscore), and (c) presence of AD based on CERAD score. These findings suggest that our graph embeddings can identify potential novel population subgroups (c1-c5), each statistically enriched with donors exhibiting AD progression pathologies and cognitive decline.

### Phenotypic population trajectories for AD progression, cognition, and neuropsychiatric symptoms

AD progression exhibits significant heterogeneity across donors, particularly in terms of age of onset, rate of cognitive decline, and worsening of disease pathology. Capturing individual-specific disease mechanisms can therefore provide insights into disease trajectories at a population level. In this study, we used donor graph embeddings, extracted from our pre-trained KG-GNN model, to infer population-level trajectories for AD phenotypes (Extended Figure 1c). By analyzing donor graph embeddings corresponding to different phenotypes, we computed phenotypic pseudotimes for these embeddings (Figure 3a, see **Methods**). Specifically, these graph embeddings allowed us to temporally order donors by assigning pseudotimes based on their phenotypes (e.g., AD, Braak, CERAD).

**Figure 3:**
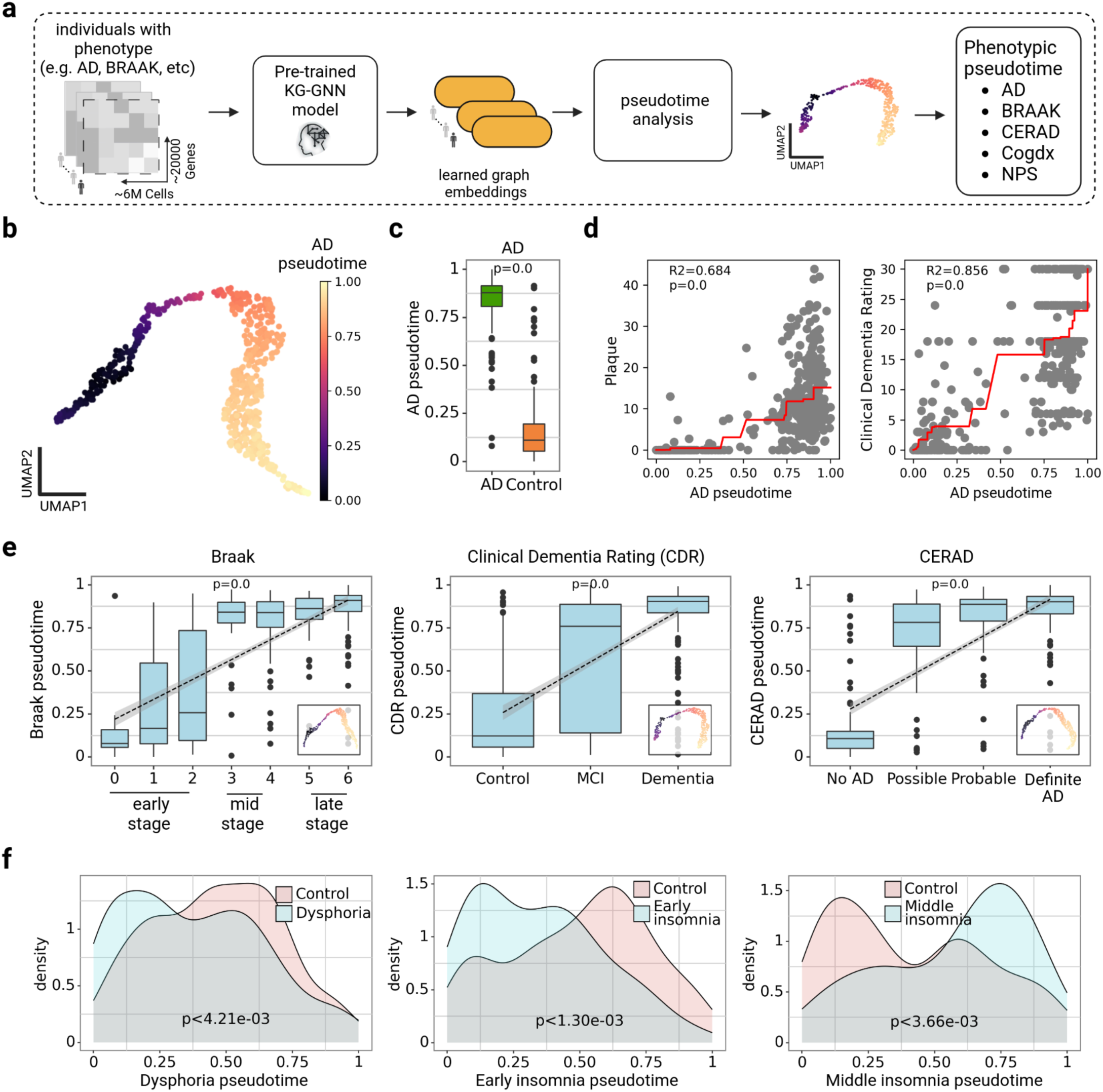
Population-level pseudotime analysis uncovers phenotypic trajectories for AD progression, cognition, and NPS using pre-trained KG-GNN model. **a,** Graph embeddings are extracted for donors with different phenotypic information using the pre-trained KG-GNN model. Then pseudotime analysis of graph embeddings is done to uncover phenotypic trajectories, **b,** AD phenotypic trajectory captured for graph embeddings of donors with AD vs. controls, **c,** Boxplot comparing pseudotimes from **b.** shows controls appearing earlier compared to donors with AD having a later occurrence, **d,** Comparison of AD trajectory pseudotimes (from **a.**) with AD pathology (Plaque) and cognition (Clinical Dementia Rating score) are consistent **e,** Boxplots showing pseudotimes computed from graph embeddings for donors with AD pathology and cognition reveal an increasing trend with stage progression; left: Braak stages=early vs. mid vs. late (Mann-Kendall, *P* = 0), middle: CDRScore=Control vs. Mild Cognitive Impairment (MCI) vs. Dementia (Mann-Kendall, *P* = 0), right: CERAD=No AD vs. Possible vs. Probable vs. Definite AD (Mann-Kendall, *P* = 0); inset plots: graph embeddings based UMAPs colored by pseudotime, **f,** Density plots showing the distribution of neuropsychiatric symptoms (NPS) across donors diagnosed with NPS including Dysphoria (*P <* 4.21e-3), Early Insomnia (*P <* 1.30e-3), and Middle Insomnia (*P <* 4.21e-3) across NPS phenotypic pseudotimes.

We first inferred a trajectory for donors with AD diagnosis versus Controls, assigning an AD pseudotime to each donor (Figure 3b). Our findings revealed that donors diagnosed with AD were assigned later pseudotimes compared to the Controls (Figure 3c). Furthermore, when correlating this AD pseudotime against AD pathology (Plaque counts) and cognition (CDRscore), we observed a clear progression trend, with increasing plaque levels and higher CDRscores associated with later pseudotimes (Figure 3d, see **Supplementary Note 3**). To further explore AD phenotypic pseudotimes, we inferred trajectories for progression and cognition phenotypes like Braak, CERAD, and CDRScore (Figure 3e). Across all three phenotypes, we observed a consistent trend of increasing pseudotime associated with more advanced stages of disease severity. This demonstrates that individual-level graph embeddings can effectively uncover phenotypic trajectories, as evidenced by the alignment of increasing AD pseudotime with more advanced stages of AD.

NPSs often co-occur in individuals with AD, with different symptoms manifesting at distinct stages of the disease^29^. For instance, dysphoria is reported to be more prevalent during the early stages of cognitive impairment^29,30^. Consistent with previous findings, our analysis revealed that pseudotimes inferred from graph embeddings of AD donors diagnosed with dysphoria were closer to zero or initial stages of AD progression (Figure 3f**: left**). Similarly, pseudotime analysis of AD donors diagnosed with early- and mid-insomnia revealed that these sleep disturbances occur along different stages as AD progresses (Figure 3f**: mid, right, Supplementary** Figure 7).

We extended this analysis to the SEA-AD cohort to independently validate the phenotypic trajectories derived from the graph embeddings generated by our model. We first constructed the *bio-diffused* PFGs for 80 donors from SEA-AD and extracted their graph embeddings using our pre-trained KG-GNN model (Extended Figure 2a, see **Methods**). We then compared the pseudotimes inferred from these embeddings with the continuous pseudoprogression score (CPS)^17^, which used a machine-learning model on quantitative neuropathology measurements and immunohistochemical stains. Interestingly, the two measures had a strong concordance, with a Pearson correlation of 0.63 (P < 1.38e-3, Extended Figure 2b). Furthermore, the pseudotimes effectively captured disease severity stages in AD progression (Thal, CERAD) and cognition (cogdx) (Extended Figure 2c-e).

### Prioritization of personalized cell type interactions, genes, and regulatory networks for AD

Graph embeddings improved phenotype prediction and provided insights into disease progression (e.g., population trajectories). Our next step was to perform a more granular analysis of the PFGs by examining the nodes and edges within each donor graph. The nodes include cell types and genes (TFs and TGs). The edges include cell type interactions and cell-type gene regulatory links (cell types to TFs, TFs to TGs). We used the attention scores from our trained KG-GNN model as the edge importance scores and designed an importance scoring metric for nodes of the PFG for each donor using the edge importance scores (see **Methods and Supplementary Note 7**). As the model was trained to classify AD vs. Control, the importance score represents the AD importance scores of each edge and node of the PFG.

As our KG-GNN model had higher AD classification performance than using gene expression alone (**Supplementary** Figure 5), we wanted to test that the importance scores from our model provide deeper insights compared to traditional gene expression based approaches. We first compared the node importance scores of different cell types with the cell type fractions across all samples. We observed that cell type fraction did not affect cell type importance (Figure 4a). Further, EN_L5_6_NP, EN*_*L6_IT_2, and IN_LAMP5_LHX6 cell types had the highest importance scores for AD pathology. When comparing with SCZ, we also identified several cell types with higher importance scores among AD donors than SCZ donors (**Supplementary** Figure 8). Using the Mann-Whitney U Test, the top three cell types with significantly higher importance scores among AD donors were IN_PVALB (FDR < 9e-4), EN_L6_CT (FDR < 2e-3), and IN_SST (FDR < 3e-4). Similarly, we compared the correlation of the gene importance scores and gene expression with clinical (Age) and pathological (CDR score, cognitive tau resilience, plaque, and polygenic risk scores) phenotypes for AD contrast. We saw higher correlations between these phenotypes and the importance scores compared to gene expression (importance: corr = 0.117; expression: corr = 0.067, Figure 4b**, Supplementary Table 2**). We observed better predictive capabilities using our importance scores among AD vs. controls for several genes that were not identified by gene expression. For example, gene expression of the GAD2 gene in IN_VIP cell type was not correlated with PRS, while importance scores identified a positive correlation among AD donors and a negative correlation among control donors (**Supplementary** Figure 9). Extending our analyses beyond AD phenotypes and PRS, we saw a higher correlation of importance scores in comparison to gene expression for SCZ, Braak, AD Resilience, and Sleep/Weight Gain/Guilt/Suicide(S/W/G/S) phenotypes (**Supplementary** Figure 10a**-f**). Thus, these observations suggest that our gene importance scores are more information-dense than gene expression and confer knowledge beyond AD pathology alone, allowing for risk prediction concerning diseases such as SCZ and clinical phenotype prediction.

**Figure 4:**
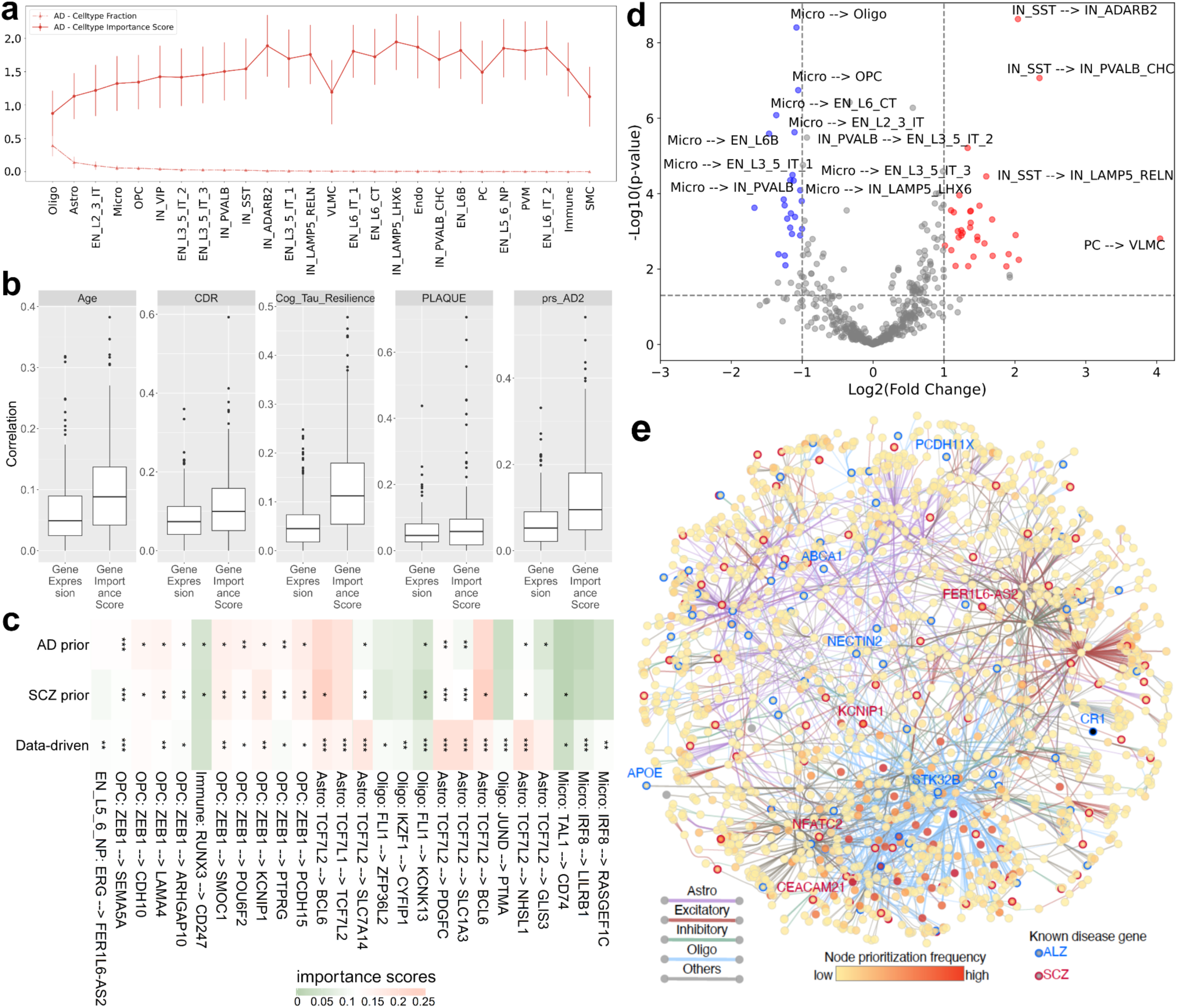
Cell type and interaction score evaluation and comparison. **a,** Identifying important cell types for AD phenotype. Dotted lines represent cell type fraction, and solid lines indicate cell type importance score. The plot is sorted by cell type fraction. **b,** Correlation comparison of importance score and gene expression for different clinical (age) and pathological phenotypes (CDR score, cognitive tau resilience, plaque count, and polygenic risk score). **c,** Variance heatmap of different gene regulatory (TF-TG) links across control and AD donors (annotated for significance) using importance scores from different biological priors for attention head groups. Green/Red: low and high difference in average importance score between control and AD donors. **d,** Volcano plot of log-fold change against significance for cell type-cell type interactions. Blue: control; red: AD. **e,** Prioritized subnetwork (top 1% edges across cell types within each donor) showcasing connectivity among cell types, TFs, and target genes for AD phenotype. Each ellipse represents a gene, shaded along a yellow to red gradient corresponding to the frequency of occurrence in prioritized nodes across donors. Edges represent regulatory links between TFs and target genes and are colored uniquely for astrocytes, oligodendrocytes, excitatory neurons, and inhibitory neurons. Edges in all other cell types are colored gray for ease of visualization. Borders of genes linked to AD and SCZ in the literature are shaded blue or red, respectively, and a few such genes are labeled with a similar color scheme.

Since our importance scores are generally more correlated with clinical phenotypes, and both gene regulatory links and cell type-cell type interactions provide insights into disease mechanisms, we compared importance scores between AD and controls for individual interactions. Our model provides three importance scores: AD-prior, SCZ-prior, and data-driven. The importance scores with each prior displayed significantly differing prioritizations, leading to uniquely prioritized edges (Figure 4c**, Supplementary** Figure 11). For example, TF *IRF8* regulatory interactions in microglia had significantly higher data-driven importance scores in controls (*IRF8*-*RASGEF1C*: P < 7e-3; *IRF8*-*LILRB1*: P < 9e-4). Changes in *IRF8*, a crucial microglial TF for AD, can cause abnormal expression of many AD-related genes^31^. Similarly, interactions from *TCF7L2* in astrocytes had higher importance scores across all priors in AD donors. *TCF7L2* is associated with the Wnt/β-catenin signaling pathway which plays a crucial role in AD^32^. *ZEB1* also plays a major role in epigenetic regulation in AD and had higher importance scores in AD donors^33^. A complete list of all the regulatory edge differences between AD and controls is available in **Supplementary Data 1**. We then examined differential importance scores on cell type interactions to identify AD-associated cell type interactions. The top three significant cell type interactions among AD donors are PC-VLMC (logFC = 4.050, P < 1.58e-03), IN_SST-IN_PVALB_CHC (logFC = 2.347, P < 9e-08), and IN_SST-IN_ADARB2 (logFC = 2.043, P < 3e-09). Similarly, the top three interactions among controls are VLMC-IN_SST (logFC = −1.673, P < 3e-04, Micro-EN_L6B (logFC = −1.467, P < 3e-06), and Micro-EN_L6_CT (logFC = −1.365, P < 9e-07). We also observed that interactions with microglia and IN_SST as source cell types were enriched in control and AD donors, respectively (Figure 4d). Microglia are known to have many effects on AD pathology and therapeutic intervention, which corroborates their high importance^34^. Thus, the importance score not only provided a comprehensive analysis of cell type importance but also facilitated the identification of significant cell type interactions, particularly highlighting the role of Microglia and IN_SST in AD.

We wanted to examine prioritized TFs and their regulatory relationships further as they pertained to specific cell types and disease classifications. We visualized and combined the top 10% of edges from each donor (based on our importance scores) into a consensus functional genomic network spanning all prioritized cell types (Figure 4e). This subnetwork illuminated several biologically relevant features of our model. For instance, visualization of the subnetwork suggests clear edge segregation into three main clusters consisting mainly of oligodendrocytes, astrocytes, and excitatory neuronal cell types, indicating that our model consistently prioritized regulatory edges in these cells across donors. We also recorded the occurrence of prioritized genes across donors and found that most frequently prioritized genes are predominantly regulated in oligodendrocytes.

Furthermore, the model prioritized several known AD and SCZ genes and illuminated their cell type-specific roles. For instance, *PCDH11X*, an aging-related gene associated with late-onset AD in GWAS^35^, is prioritized in excitatory neurons in our model. A previous study identified variants in the *CR1* gene associated with AD^36^. Our model suggests that *FOSB* mediates *CR1* regulation, and this regulation is relevant for AD PVM cells. We also observed a considerable representation of known SCZ genes within this subnetwork. Overall, our model effectively identifies and prioritizes a subnetwork that recapitulates known AD biology, particularly regarding cell-type-specific regulation of several genes with prior evidence. Thus, other genes in the prioritized subnetwork are likely candidates that have remained elusive thus far.

### Personalized prioritization enables subpopulation-level analysis in diseases and clinical phenotypes

As our importance scores are differentiable across disease and control groups, we next sought to compare cell type interactions and regulatory links across diseases and clinical phenotypes. We extended our analysis to other phenotypes, including cross disorders (AD vs SCZ), AD progression phenotypes, and ancestry. We first looked at differential importance scores for these phenotypes. Several TFs from inhibitory neurons (IN) and excitatory neurons (EN) were enriched in SCZ compared to AD, and TFs from astrocytes were enriched in AD donors (Figure 5a**, Supplementary Data 2**). Specifically, we identified several known SCZ TFs like *TCF4*^37^, *BCL11A*^38^, *RBFOX2*^39^, and *ZMAT4*^40^ as the top enriched genes in SCZ donors which were also significantly differentially expressed in a study of 388 human brains^41^. Similarly, for Braak stages, we identified several TFs from IN cell types enriched in early Braak stages while TFs from astrocytes and microglia cell types enriched in late Braak stages (Figure 5a). Using the differential gene importance scores, we were able to identify cell type and TF relationships across diseases and disease progression.

**Figure 5:**
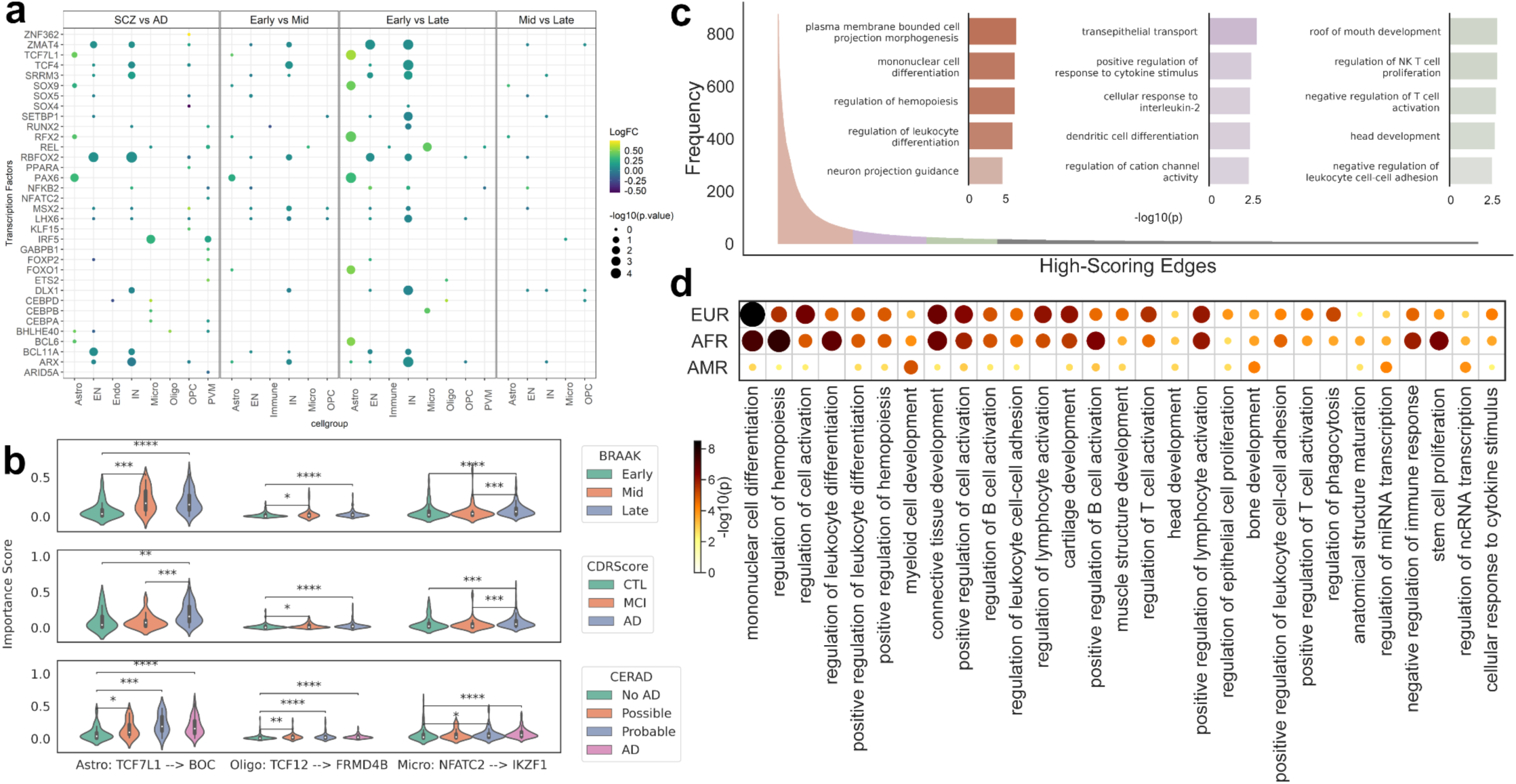
Cross disorder and progression prioritization using our personalized importance scores. **a,** Differential gene importance scores between cross disorder (AD vs SCZ) and AD progression (early vs mid vs late Braak). The x-axis is different cell types, and the y-axis represents the transcription factors. Each dot varies by size and color. Light green denotes positive log fold change and darker green denotes negative fold change. The dot size indicates the significance of the differential analysis (-log10(p-value)) **b,** Edge importance distributions for select gene regulatory links, compared for various AD progression phenotypes and annotated with ANOVA. **c,** Histogram of edges scoring in the top 5% of data-driven importance scores among the whole population and GO-term enrichments (bar plots) for genes in the resultant highly conserved (top 2%, 51-875 occurrences, orange), conserved (top 2-4%, 23-51 occurrences, purple), and unique (top 4-6%, 15-23 occurrences, green) edge groups. Only the top 20% of edges are shown in the histogram for visualization purposes. **d,** Enrichment of highly conserved edges from African (AFR, n = 120), admixed American (AMR, n = 99), and European (EUR, n = 792) subpopulations.

To further examine disease relationships with importance scores, we analyzed edge importance score distribution differences across phenotypes, particularly Braak stages, CDR score, and CERAD score (Figure 5b). We found several significant relationships, including the microglia edge *NFATc2*-*IKZF1*, which displays significant differences in distribution across all Braak stages (early vs late: P < 1.79e-6). *NFATc2* has been linked to AD pathology in mouse models through modulation of microglial activation^42^. Additionally, we found the *TCF7L1*-*BOC* edge in astrocytes to separate extreme cases of CERAD scores (No AD vs AD: P < 2.31e-5). *TCF7L1* is known to be a highly astrocyte-specific regulator in AD pathology^43^. Our model was able to effectively uncover significant prioritization differences across AD progression and pathology phenotypes, revealing potentially novel disease regulatory relationships.

We extended our analysis beyond disease phenotypes to understand conservation of regulatory links across the dataset population. As the population includes donors with different disease phenotypes beyond AD, we used data-driven importance scores rather than AD-prior importance scores. As shown in Figure 5c, most of the edges are present in few donors, but some select edges are shown to be common in PFG graphs. These edges may be separated into groups based on frequency then enriched to examine patterns within the common edges. For this analysis, we sorted edges in the top 5% of data-driven importance scores by frequency and performed enrichments on groupings of the constituent top genes. We grouped these edges into three categories: highly conserved (top 2%), conserved (top 2-4%), and unique (top 4-6%) edges based on empirical observation. Each edge was split into constituent genes to construct a gene list for enrichment (Figure 5c, see **Methods**). Among the enriched terms in the highly conserved category, we observed relationships with regulation of leukocyte differentiation. Low leukocyte counts are associated with increased risk for developing AD^44^. We also found dendritic cell differentiation to be enriched among conserved edges. Dendritic cells have been shown to be associated with AD progression and depression^45^. Interestingly, the head development pathway was also significantly enriched among unique edges. The model allows for intuitive prioritization of regulatory links and genes, which can be used for further downstream analysis.

Differing subpopulations, such as ancestries, have been shown to have significant AD risk disparities^46,47^. We leveraged our data-driven importance scores across ancestries to complement the growing body of research which finds racial disparities in AD risk, performing enrichment of highly conserved genes for individual ancestries (Figure 5d). In particular, we analyzed African (AFR, n = 120), admixed American (AMR, n = 99), and European (EUR, n = 792) populations. Regulation of phagocytosis had the highest significance among AFR (AFR: P < 6.31e-6; AMR: P < 2.52e-4; EUR: P = 1). Phagocytosis of amyloid-beta (Aβ) plaques, particularly by microglia, is strongly related to AD severity^48^. The analysis also reveals obscure and previously unseen relationships. For instance, connective tissue development was particularly significant across AFR and admixed AMR populations (AFR: P < 2.00e-7; AMR: P < 2.00e-7; EUR: P < 3.17e-3). Expression of connective tissue growth factor is a regulator of Aβ plaque, which directly pertains to AD pathology^49^. We also found several common enrichments, including Myeloid cell development (AFR: P < 6.31e-4; AMR: P = 1.00e-4; EUR: P = 1.00e-5) which is generally associated with AD pathology^50^. Ancestry analyses for AD and SCZ-prior-based importance score distributions are available in **Supplementary** Figure 12. The edge importance scores revealed shared and unique pathways among subpopulations such as ancestry, offering a more detailed understanding of the mechanisms underlying AD pathology.

### Genotype association and gene regulatory QTL (grQTL) analysis

Genetic variation influences gene regulation through various mechanisms like alterations in cis-regulatory elements, which leads to cell type differences in gene expression, impacting disease susceptibility or progression^41,51–53^. We first explored the relationship between PRS and importance scores derived from iBrainMap to understand association between disease heritability and our PFGs (Figure 6a). We identified several edges correlated with PRS scores. The top correlating edge consisted of the genes BCL6 and CDH23 (corr = −0.861, P < 3.26e-4), which are known to be linked with AD^54,55^. Additionally, the second edge links Immune cells and SPI1 (corr = −0.811, P < 4.41e-3), which has been associated with AD^56^ and the age of Alzheimer’s onset^57^. *BATF* in EN_L3_5_IT_1 cell type has a high correlation with SCZ PRS (corr = 0.862, P < 1.36e-3). *BATF* has been associated with immune dysfunction in SCZ^58^. Similarly, IN_LAMP5_RELN to *ARID5B* edge (corr = −0.911, P < 1.68e-4) was highly correlated with the SCZ PRS measure.

**Figure 6:**
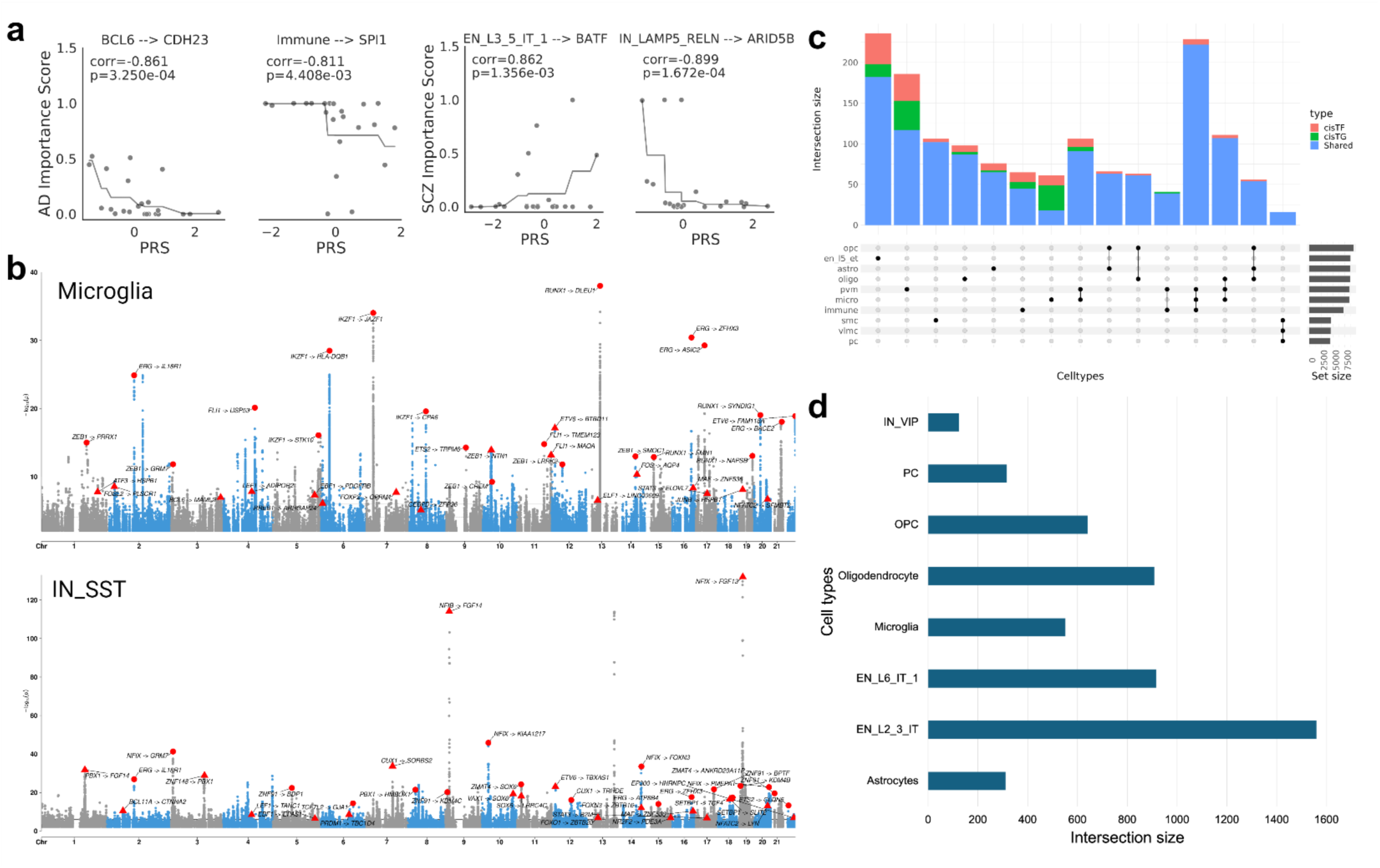
genotype association with gene regulatory networks. **a,** Correlation between polygenic risk score and importance score for each regulatory link (Transcription Factor to target gene). The X-axis indicates the polygenic risk score and the y-axis represents the importance score. AD importance score is showcased on the left and SCZ importance score on the right. Each dot in the plot represents a donor. The gray line is a monotonically increasing/decreasing best-fit line, included for clarity. **b,** Manhattan plot showing grQTLs for microglia (top) and IN_SST (bottom) cell types. The X-axis indicates the chromosome locations, and the y-axis represents the association significance calculated using -log10(p-value). Triangles indicate the cisTF-grQTL meaning the SNP is associated with the TF coordinate, while circles represent cisTG-grQTL. **c,** Variation of the number of cell type grQTLs per cell type. Blue indicates shared grQtls between cisTF and cisTG, orange indicates cisTF-grQTLs, and green indicates cisTG-grQTLs. **d,** grQTLs intersected with psychEncode2 (link) eQTLs across different cell types.

As the edge correlation with PRS scores was high, we wanted to understand how genetic variants affect gene regulation in different brain cell types. Therefore, we performed genotype association with cell type TF-TG regulatory links. We specifically tested the association of genotypes of SNPs within a 1-Mb cis region (including the gene body) of both TF and TG genes with the TF-TG regulatory score from SCENIC for 27 cell types (see **Methods**). We call the variants associated with the TF-region cisTF-grQTL and variants associated with the target gene region cisTG-grQTL. In total, we identified 40,052,033 grQTLs across 27 cell types (FDR < 1e-5, Fig. 6b). Among these ∼40M grQTLs, there were 9,733,663 cisTF-grQTLs and 30,318,370 cisTG-grQTL. On average, we identified 1,483,408 grQTLs per cell type with microglia having one of the lowest counts (1,003,126 grQTLs) and IN_SST having one of the highest counts (1,718,902 grQTLs). The grQTL plots for all the remaining cell types are available in **Supplementary Data 3.**

The number of independent grQTLs varied across different cell types with 274 grQTLs identified in EN_L5_ET cell type and PVM cell type had the lowest count with 57 (Figure 6c). The majority of cell types shared the grQTLs. Furthermore, most of the grQTLs were shared between cisTF and cisTG, with some exceptions. For example, Microglia has more cisTG-grQTLs than cisTF-grQTLs and shared grQTLs. We then compared the grQTLs with recent brain cell-type eQTLs^41^ and found several cell types with high intersection of the SNP-gene pairs (e.g., n = 1,559 for EN_L2_3_IT, n = 916 for EN_L6_IT_1, and n = 918 for oligodendrocytes, Figure 6d). Our analysis revealed significant associations between genetic variants and cell type specific gene regulation in the brain, identifying grQTLs for 27 cell types. This provides novel insights into how genetic variation influences gene regulation in different brain cell types, potentially contributing to the heterogeneity observed in AD and other neuropsychiatric disorders.

Next, we used genotype data to impute graph embeddings for new donors for whom only genotype data was available (Extended Figure 3a). With these imputed graph embeddings, we classified donors into different disease phenotype groups (see **Supplementary Note 8**). For instance, using genotype information from donors in the ROSMAP^59^ cohort, we imputed their graph embeddings and classified them into Early vs. Late Braak stages, achieving an AUC score of 0.57 (Extended Figure 3b). We then clustered the donor graph embeddings to identify potential novel subtypes (Extended Figure 3c), ultimately inferring seven clusters. We further performed hypergeometric tests on these clusters to identify novel subtypes (Extended Figure 3d). In particular, we found that clusters c2-c6 were enriched with CERAD stages, which increase in severity from ‘No AD’ to ‘Definite’. By leveraging genotype data to impute graph embeddings, we were able to predict disease phenotypes and identify novel subtypes in AD.

## Discussion

In this study, we present what is, to our knowledge, the first personalized functional genomics (PFG) analysis of AD using population-scale snRNA-seq data. Through the iBrainMap framework, we constructed PFGs for 1,494 donors in the PsychAD cohort, capturing cell type interactions and gene regulatory mechanisms at donor-level. We applied graph-based learning on these PFGs to generate graph embeddings, which were used to classify phenotypes, stratify donors, and reveal potential novel subgroups. Additionally, the model produced personalized importance scores for genes, cell types, and interactions, prioritizing personalized subnetworks, including those involving cell type interactions and gene regulation for various phenotypes. We also computed cell type gene regulatory QTLs (grQTLs) to associate genotypes with cell type-specific gene regulation. Our findings were validated on external snRNA-seq datasets, including SEA-AD and ROSMAP, demonstrating the framework’s utility in phenotype classification and identification of personalized phenotypic subnetworks. These results have been compiled into a personalized functional genomic atlas for Alzheimer’s disease, which includes PFGs, a pre-trained knowledge-guided graph neural network (KG-GNN) model for phenotype prediction and functional genomic prioritization, and cell type-level grQTLs associating SNPs with gene regulatory mechanisms. This atlas is accessible via an interactive Shiny web app, and the iBrainMap framework and pre-trained model are available as open-source tools on GitHub.

The robustness of our KG-GNN model was further confirmed through independent validation on the SEA-AD dataset, despite lacking some cell types and genes present in the training data from the MSSM cohort. By employing a sub sampling technique, our model successfully captured and processed information from incomplete data, maintaining its predictive power and analytical capabilities even with datasets differing in composition from the training data. This flexibility in handling incomplete PFGs not only underscores the model’s adaptability but also broadens its applicability across diverse datasets, highlighting its effectiveness as a tool for personalized functional genomics analysis in AD and potentially other complex disorders.

Genetic variability in complex diseases like Alzheimer’s enables the study of personalized functional genomics. By capturing this variability, our approach can drive the development of tailored therapeutic strategies. Prior research has shown that cell type-specific networks can enhance the prediction of potential novel drug targets and AD-related genes, which is valuable in drug repurposing and predicting clinical phenotypes of AD^61^. Additionally, a computational drug-repurposing screen identified bumetanide as a potential treatment to lower AD risk in donors carrying the APOE ε4 variant^62^. Our study offers new insights into potential biomarkers and genetic regulatory mechanisms that can be targeted for more personalized and effective treatments, further contributing to the field of precision medicine. Moreover, our personalized functional genomics approach can be further extended to other complex diseases exhibiting significant genetic variability like schizophrenia, bipolar disorder, autism spectrum disorder^41^ and cancer^63^, especially when population functional genomics data is available.

Our work has several limitations. First, the personalized functional genomics graphs are created by selecting key transcription factors and target genes for each cell type of an individual, focusing on significant regulatory mechanisms. While this approach highlights meaningful interactions and our model performs well on independent datasets, it only captures only a portion of the regulatory activity. Future work could incorporate scaling the personalized functional genomics graphs (e.g., TFs, TGs) to provide a complete representation of an individual’s regulatory mechanisms.

Additionally, our graph neural network model treats all nodes and edges in these graphs as uniform, which overlooks cellular and molecular networks’ inherent diversity and complexities. To address this, future efforts could explore using more sophisticated models, such as heterogeneous graph neural networks^64^, that can distinguish different node and edge types, including various cell types, genes, and regulatory mechanisms. Moreover, extending our analyses to emerging single-cell multimodal such as epigenomics, proteomics, and spatial data, could provide a more holistic view of disease mechanisms.

## Methods

### Datasets and data preprocessing

#### Dataset

Our analysis is focused on the population-level snRNA-seq data from the PsychAD consortium covering the dorsolateral prefrontal cortex (DLPFC) brain region from 1494 donors, derived from three cohorts: NIH NeuroBioBank at the Mount Sinai Brain Bank/MSSM (M), Rush Alzheimer’s Disease Center/RADC (R), and the NIMH-IRP Human Brain Collection Core/HBCC (H). The PsychAD data description is available in the “PsychAD dataset” (See Supplementary Information). Our analyses utilized disease diagnosis (e.g. AD, SCZ, DLBD), quantitative assessment of the disease stages for donors with AD (e.g. Braak or CERAD), and diagnosis of neuropsychiatric symptoms (e.g. depression or weight loss). Accordingly, these donors were categorized into three levels: disease versus control (AD, SCZ, DLBD, AD Resilience), AD progression (Braak stages, Cogdx, CERAD), and NPSs (Dysphoria, DecInt (Anhedonia), Sleep/Weight Gain/Guilt/Suicide (S/W/G/S), Depression/Mood (D/M)) (Extended Figure 1c**, Supplementary Note 3**). We mainly used the MSSM (M) cohort for all the training and used RADC (R) as well as an external dataset from The Seattle Alzheimer’s Disease Brain Cell Atlas/SEA-AD (S)^18^ for independent validation.

#### Data processing and feature selection

PsychAD snRNA-seq data was preprocessed as described in Lee et al.^19^ and Fullard et al.^20^. The preprocessing involved identifying 5,000 highly variable genes (HVGs). We selected the protein-coding genes from these 5,000 HVGs and intersected them with the genes from the SEA-AD dataset. We ended up with 2,766 HVGs which were used as features in our analysis. We applied standard Scanpy (v1.9.3) functions to preprocess SEA-AD data. For SEA-AD, we selected DLPFC regions of 39 donors with dementia and 39 without dementia to retain individual balance. For independent validation, we constructed *bio-diffused* PFGs for the donors and included the same set of 2,766 HVG genes as features as the PscyhAD dataset.

#### Construction of personalized functional genomics graph

For each donor, we define his/her Personalized Functional Genomics graph (PFG) as a directed graph with two major node types: cell types and genes where gene nodes include cell type-specific transcription factor (TFs) and target genes (TGs) nodes. Each edge in the PFG conveys distinct information depending on the type of nodes. For example, cell type-cell type edge captures intercellular relationships and communications, TG-TG links capture gene regulatory relationships, and cell type-TF links capture cell type specificity relationship of TFs. As some of the donors had fewer cells for the cell type interactions and gene regulatory relationship algorithms to capture (**Supplementary** Figure 13), we were able to construct PFGs for 1478 out of 1494 donors. Below, we provide details on constructing each component of PFG.

#### Inferring cell type interactions

For each donor, the cell type to cell type links were constructed using CellChat^65^ to quantify the intercellular interactions. CellChat provides a consolidated score for each cell type to cell type interaction by adding all the probabilities of ligand-receptor interactions between two cell types. We used snRNA-seq data from each donor as input to CellChat with 27 subclass information as the label. We used the default settings in CellChat to extract the scores using the function computeCommunProb. We then normalized the scores by cell type and retained all links with the normalized score above a 0.5 threshold to construct cell type to cell type links for each donor.

#### Extracting cell type gene regulatory links

We first used GRNBoost2 on the snRNA-seq data for each donor to deduce gene co-expression networks across all cell types. SCENIC^66^ was then used to infer cell type gene regulatory links. Specifically, we applied runscENIC_1_coexNetwork2modules and runscENIC_2_createRegulons functions in Python with default setting (constraining the TF search to 10-kbps radius around the TSS or 500 bp upstream) to identify regulons. Regulons with at least 10 genes were scored in each cell using runscENIC_3_scoreCells. We then computed AUCell enrichment based on the top 1% of genes detected per cell. We incorporated the regulon specificity score (RSS)^67^ to evaluate the cell type specificity of the regulon. For each cell type, we select the top 10% of the regulons based on the RSS score as the cell type regulons. We then keep the top 10% of the TGs for each cell type regulon. Together, these comprise the cell type gene regulatory links for each donor.

#### Definition of PFGs

Let *G*_i_ = (*V*_i_, *E*_i_) (Extended Figure 1a) be a PFG for donor *i*, with *N*_i_ = |*V*_i_| nodes and *E*_!_ representing the edges. Then, an edge *e*_i,st_ ∈ *E*_i_ connecting the source node *s* to a target node *t* represents one of the following relationships:

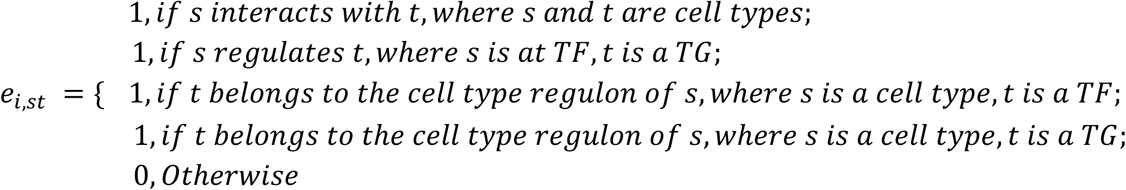

The node features used for each PFG depend on the type of node. We used the average gene expression of 2,766 highly variable genes (HVGs) from all cells as the features for cell type (CT) nodes and the coexpression of these HVGs with the gene expression for the TF/TG nodes.

### Integrating prior biological knowledge into PFGs via network diffusion

We then applied network diffusion to propagate the influence of known disease genes on the PFGs (see **Supplementary Note 2**). This diffusion process allows us to incorporate known disease-related information into our graph structure, potentially enhancing the model’s ability to capture disease-relevant patterns. We currently use well-known disease genes for AD and SCZ extracted from the DisGeNet^27^ database and filtered based on manually curated sources with “CTD_human” or “GWASCAT” identifiers. Additionally, we downloaded high-confidence SCZ genes from the PsychENCODE project^28^. In total, we identified 361 AD genes and 945 SCZ genes. The resulting matrices are called “bio-diffused PFGs” and are used to train our graph neural network model. While we focus on AD and SCZ in this paper, this method can be applied to any gene-of-interest (GOI) list.

### Graph classification using Knowledge-guided Graph Neural Network (KG-GNN)

The KG-GNN model is based on the multi-head Graph Attention Networks (GAT)^68^ that learns from arbitrary *bio-diffused* PFGs as inputs for the binary graph classification of healthy versus AD. It is an interpretable machine learning model that integrates prior biological knowledge, personalized functional genomics, and biologically driven multi-head graph attention networks to derive insights about disease and underlying disease mechanisms at an individual level. The details of architecture, training procedure, validation, and benchmarking are provided in **Supplementary Note 1.** The input to the KG-GNN mode is the PFGs. The trained model outputs graph embeddings and personalized AD importance scores for nodes and edges.

### Population subtyping and enrichment based on graph embeddings

We clustered graph embeddings extracted for donors with AD vs. Controls using the function sc.tl.louvain() from the Python package scanpy^69^. Before clustering, we identified the nearest neighbors using the sc.pp.neighbors() function and computed the umap embeddings using the sc.tl.umap() function. Then we performed Louvain clustering using the function sc.tl.louvain() and set the resolution = 0.35 to get 5 clusters. We clustered the graph embeddings to identify potential novel subtypes. To identify novel subtypes, we performed enrichment analysis using a hypergeometric test to see if any clusters were enriched with AD progression stages. We used the stats.hypergeom.cdf() from the Python package Scipy^70^ for this analysis and reported the -log10(p-value) of phenotype enrichment in each cluster.

### Phenotypic population trajectory inference

We inferred phenotypic trajectories across AD phenotypes based on individual graph embeddings extracted from our pre-trained KG-GNN model. Here we used a diffusion-based algorithm^71^ within the scanpy package in Python to infer trajectories. Given a phenotype, we run the functions sc.tl.diffmap() and sc.tl.dpt() for individual graph embeddings associated with the phenotype. We annotate a root individual based on the phenotype. For example, we set a randomly selected individual with Braak stage 0 as the root for comparing AD vs. controls.

Similarly, for other AD progression phenotypes, we set the root as follows: (1) Braak pseudotime: randomly selected a donor within Braak stage 0 (2) CERAD pseudotime: randomly selected a donor within CERAD stage 0/No AD, (3) Cogdx pseudotime: randomly selected a donor with cognitive diagnosis set to 1/Control. Next, to infer the pseudotimes for different NPS, we again set a randomly selected a donor with a CERAD score of 2 as the root. This is because all donors diagnosed with NPS have a CERAD score equal to 2 at least.

To infer the trajectory for SEA-AD donors, we first extracted their graph embeddings from the pre-trained KG-GNN model. Then using similar functions as above (sc.tl.diffmap(), sc.tl.dpt()), we computed a trajectory by randomly selecting a donor with CERAD score = “Absent” as the root.

### Calculation of importance scores

The trained KG-GNN model outputs edge attentions which are used as edge importance scores. We get three different edge importance scores: AD-prior, SCZ-prior, and data-driven based on prior biological knowledge from known AD-genes and SCZ-genes. We derive the node importance scores using the edge importance scores. We use a combination of both incoming and outgoing edge importance as the basis to calculate these scores. Further details about node importance calculation are available in **Supplementary Note 7**.

### Subpopulation edge conservation based on importance scores

To determine gene conservation across donors, we first counted instances where each edge was scored in the 95th percentile or above based on the attention scores of the selected head. We then sorted the edge by frequency and established three groups: Highly conserved edges (top 2%), conserved edges (top 2-4%), and unique edges (top 4-6%). The genes associated with each edge group were compiled into lists for subsequent enrichment analysis. The grouping of the edges was based on heuristics, as the frequency with which highly prioritized edges appeared in the PFG graphs declined rapidly from the 100th to 94th percentiles.

### Gene set enrichment analysis

We used Metascape^72^ for gene set enrichment. All genes within our analysis were used as the background for the enrichment analysis. P-values were transformed into a readable format and colored according to their significance and group.

### Polygenic Risk Score (PRS) calculation

We used AD GWAS^73^ and SCZ GWAS^74^ to compute the PRS scores within the paper. Both were calculated for donors using PRS-CS^75^ and PLINK 2.0^76^. The PRS-CS-auto method was utilized, which employs continuous shrinkage priors to refine effect sizes derived from summary statistics. We used an LD reference panel from the creators of PRS-CS, based on the 1000 Genomes Project data^77^ (available at https://github.com/getian107/PRScs). Default settings were maintained for PRS-CS, including the γ-γ prior parameters *a* = 1 and *b* = 0.5, 1,000 MCMC iterations, 500 burn-in iterations, and a thinning factor of 5. The global shrinkage parameter φ was estimated using a fully Bayesian approach. PRS calculations at the donor level were performed using PLINK 2.0.

### Gene-regulation QTL (grQTL) analysis

We associated genotype with the TF-TG edge scores per cell type and called it grQTLs. To do this, we mapped cis-eQTLs within a 1-Mb window region of the TSS of both TFs and TGs using QTLtools^78^. We call the SNPs associated with the TF region cisTF-grQTL and the TG region cisTG-grQTL. We used the first three genotyping PCs, age, gender, and 50 PEER factors as covariates. Using the associated grQTLs, we then performed FDR correction and used 1e-5 as the cut-off to identify the most significant cell type-specific cisTF-grQTLs and cisTG-grQTLs.

## Data Availability

All data produced in the present work are contained in the manuscript.

## Acknowledgments

We would like to express our deep gratitude to the patients and their families who generously donated the invaluable biological material essential for the success of this study. We are profoundly indebted to their participation and commitment to advancing scientific knowledge and improving human health. We acknowledge the National Institutes of Health grants, R01AG067025 (to P.R. and D.W.), RF1MH128695 (to D.W.), R21NS127432 (to D.W.), R21NS128761 (to D.W.), U01MH116492 (to D.W.), U01MH116442 (to P.R.), R01MH110921 (to P.R.), R01MH109677 (to P.R.), P50HD105353 (to Waisman Center), National Science Foundation Career Award 2144475 (to D.W.), Simons Foundation Autism Research Initiative pilot grant 971316 (to D.W.), and the start-up funding for D.W. from the Office of the Vice Chancellor for Research and Graduate Education at the University of Wisconsin–Madison. The funders had no role in study design, data collection and analysis, decision to publish, or manuscript preparation.

## Author contributions

Conceptualization: DW, PR Methodology: PBC, SA, TJ, DW Software: PBC, SA, TJ

Formal analysis: PBC, SA, NK, TJ, DW

Investigation: XH, KG, GV, JB, JFF, DL, GEH, PR

Resources: DL, JB, KG, GV, GEH, JFF, PR

Data Curation: DL, JB, KG, GV, GEH, JFF

Writing: PBC, SA, NK, TJ, XH, DW, PR with support from all co-authors.

Visualization: SA, PBC, NK, TJ, CG, RB

Supervision: DW, PR Funding acquisition: DW, PR

All authors read and approved the final draft of the paper.

## Competing interests declaration

The authors declare no competing interests.

## Materials & Correspondence

Correspondence to Panos Roussos or Daifeng Wang.

## Code availability

The iBrainMap framework and the pre-trained model, which takes personalized snRNA-seq data as input for classifying AD and generating embeddings and importance scores, together with tutorials and demos, can be accessed at https://github.com/daifengwanglab/iBrainMap. The personalized functional genomic graphs, gene-regulation QTLs (grQTLs), and all the results from our analysis can be interactively visualized at https://daifengwanglab.shinyapps.io/iBrainMap/. All code and data used for generating figures can be accessed at https://zenodo.org/records/13635676. All graphs were visualized using cytoscape^79^ and panels of all figures were combined using BioRendr (https://www.biorender.com/).

## Supplementary Information

PsychAD dataset

PsychAD dataset.pdf

Supplementary Materials

Supplementary Notes 1-8, Supplementary Figures 1-13, and Supplementary Tables 1-2.

## Supplementary Data

**Supplementary Data 1.** The importance scores of cell type TF-TG regulatory links for AD vs. Controls. Each row is a gene regulatory link and the columns represent the importance scores and contains four scores based on prior biological knowledge (AD-prior, SCZ-prior, data-driven, and combined).

**Supplementary Data 2**. Differential TFs based on importance score across multiple phenotypes. Each row is a TF gene and the columns are cell type, p-value, fdr, log fold change (logFC), and phenotype.

**Supplementary Data 3**. Top 100 Cell type cisTF- and cisTG-grQTLs for 25 cell subclasses. The columns indicate SNP id, gene regulatory link, chromosome, position, p-value, cell type, cis type.

## PsychAD Consortium Authors

Aram Hong (1, 4, 6, 7); Athan Z. Li (10, 12); Biao Zeng (1, 4, 6, 7); Chenfeng He (9, 12); Chirag Gupta (9, 12); Christian Porras (1, 4, 6, 7); Clara Casey (1, 4, 6, 7); Colleen A. McClung (18); Collin Spencer (1, 4, 6, 7); Daifeng Wang (9, 10, 12); David A. Bennett (19); David Burstein (1, 2, 4, 6, 7, 8); Deepika Mathur (1, 4, 6, 7); Donghoon Lee (1, 4, 6, 7); Fotios Tsetsos (1, 2, 4, 6, 7); Gabriel E. Hoffman (1, 2, 4, 6, 7, 8); Genadi Ryan (13, 17); Georgios Voloudakis (1, 2, 3, 4, 6, 7, 8); Hui Yang (1, 4, 6, 7); Jaroslav Bendl (1, 4, 6, 7); Jerome J. Choi (11, 12); John F. Fullard (1, 4, 6, 7); Kalpana H. Arachchilage (9, 12); Karen Therrien (1, 4, 6, 7); Kiran Girdhar (1, 4, 6, 7); Lars J. Jensen (21); Lisa L. Barnes (19); Logan C. Dumitrescu (22, 23); Lyra Sheu (1, 4, 6, 7); Madeline R. Scott (18); Marcela Alvia (1, 4, 6, 7); Marios Anyfantakis (1, 4, 6, 7); Maxim Signaevsky (6, 7); Mikaela Koutrouli (1, 4, 6, 7, 21); Milos Pjanic (1, 4, 6, 7); Monika Ahirwar (13, 17); Nicolas Y. Masse (1, 4, 6, 7); Noah Cohen Kalafut (10, 12); Panos Roussos (1, 2, 4, 6, 7, 8); Pavan K. Auluck (20); Pavel Katsel (6); Pengfei Dong (1, 4, 6, 7); Pramod B. Chandrashekar (9, 12); Prashant N.M. (1, 4, 6, 7); Rachel Bercovitch (1, 4, 6, 7); Roman Kosoy (1, 4, 6, 7); Sanan Venkatesh (1, 2, 4, 6, 7); Saniya Khullar (9, 12); Sayali A. Alatkar (10, 12); Seon Kinrot (1, 4, 6, 7); Stathis Argyriou (1, 4, 6, 7); Stefano Marenco (20); Steven Finkbeiner (13, 14, 15, 16, 17); Steven P. Kleopoulos (1, 4, 6, 7); Tereza Clarence (1, 4, 6, 7); Timothy J. Hohman (22, 23); Ting Jin (9, 12); Vahram Haroutunian (5, 6, 7, 8); Vivek G. Ramaswamy (13, 17); Xiang Huang (12); Xinyi Wang (1, 4, 6, 7); Zhenyi Wu (1, 4, 6, 7); Zhiping Shao (1, 4, 6, 7)

## PsychAD Consortium Affiliations

1. Center for Disease Neurogenomics, Icahn School of Medicine at Mount Sinai, New York, NY, USA
2. Center for Precision Medicine and Translational Therapeutics, James J. Peters VA Medical Center, Bronx, NY, USA
3. Department of Artificial Intelligence and Human Health, Icahn School of Medicine at Mount Sinai, New York, NY, USA
4. Department of Genetics and Genomic Sciences, Icahn School of Medicine at Mount Sinai, New York, NY, USA
5. Department of Neuroscience, Icahn School of Medicine at Mount Sinai, New York, NY, USA
6. Department of Psychiatry, Icahn School of Medicine at Mount Sinai, New York, NY, USA
7. Friedman Brain Institute, Icahn School of Medicine at Mount Sinai, New York, NY, USA
8. Mental Illness Research, Education and Clinical Center VISN2, James J. Peters VA Medical Center, Bronx, NY, USA
9. Department of Biostatistics and Medical Informatics, University of Wisconsin-Madison, Madison, WI, USA
10. Department of Computer Sciences, University of Wisconsin-Madison, Madison, WI, USA
11. Department of Population Health Sciences, University of Wisconsin-Madison, Madison, WI, USA
12. Waisman Center, University of Wisconsin-Madison, Madison, WI, USA
13. Center for Systems and Therapeutics, Gladstone Institutes, San Francisco, CA, USA
14. Department of Neurology, University of California San Francisco, San Francisco, CA, USA
15. Department of Physiology, University of California San Francisco, San Francisco, CA, USA
16. Neuroscience and Biomedical Sciences Graduate Programs, University of California San Francisco, San Francisco, CA, USA
17. Taube/Koret Center for Neurodegenerative Disease Research, Gladstone Institutes, San Francisco, CA, USA
18. Department of Psychiatry, University of Pittsburgh School of Medicine, Pittsburgh, PA, USA
19. Rush Alzheimer’s Disease Center and Department of Neurological Sciences, Rush University Medical Center, Chicago, IL, USA
20. Human Brain Collection Core, National Institute of Mental Health-Intramural Research Program, Bethesda, MD, USA
21. Novo Nordisk Foundation Center for Protein Research, Faculty of Health and Medical Sciences, University of Copenhagen, Copenhagen, Denmark
22. Vanderbilt Genetics Institute, Vanderbilt University Medical Center, Nashville, TN, USA
23. Vanderbilt Memory & Alzheimer’s Center, Vanderbilt University Medical Center, Nashville, TN, USA

---

**Extended Figure 1:**
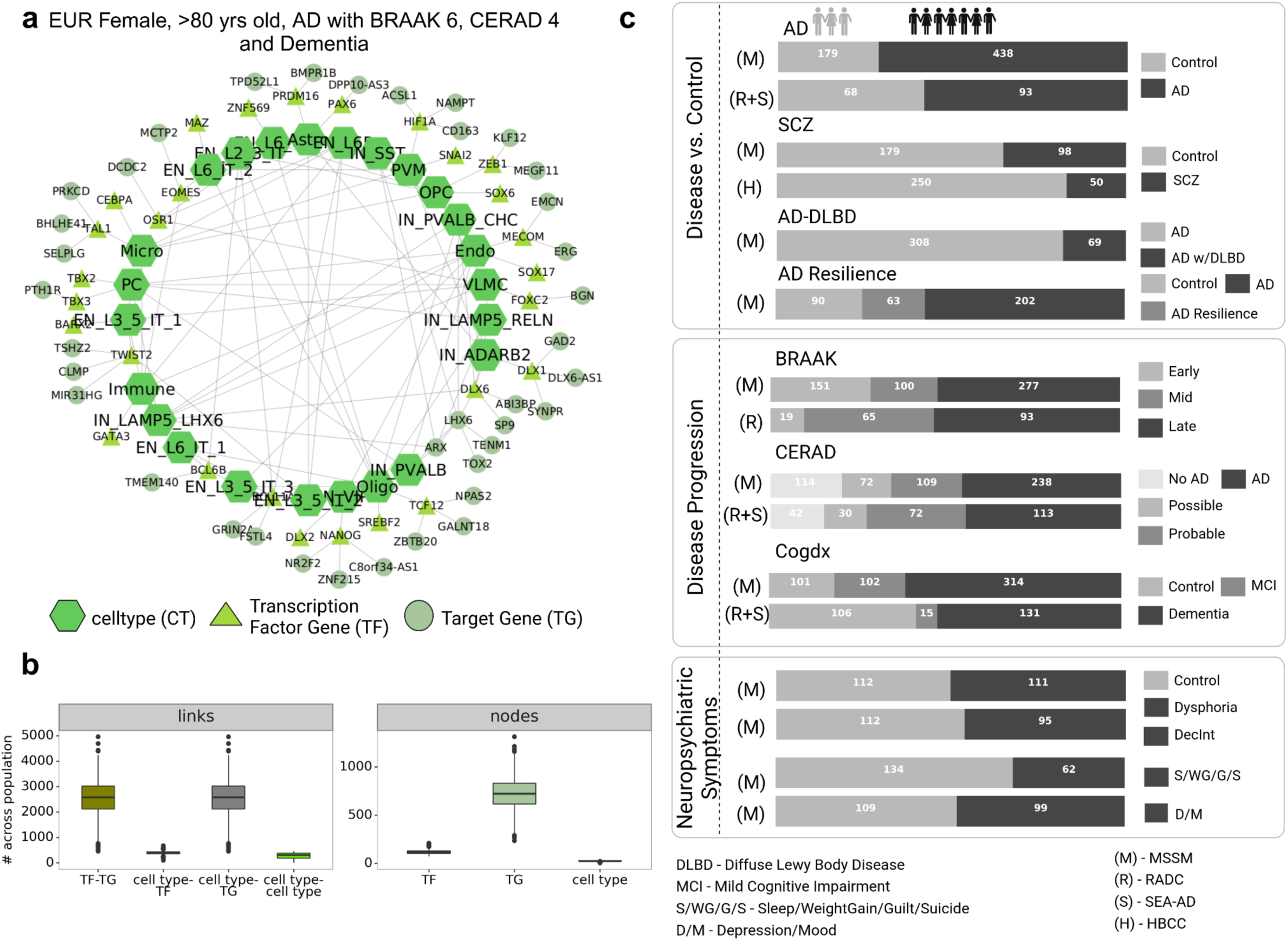
Multi-cohort snRNA-seq data for personalized functional genomic analyses. **a,** Part of a personalized functional genomics graph (PFG) for a donor from the Mount Sinai Brain Bank/MSSM (M) cohort. The donor is female with European ancestry, age >80 years old, diagnosed with AD, showing pathology for Braak stage 6 and CERAD score 4 (definite AD), and diagnosed with Dementia (see **Supplementary Note 3**). Nodes are either cell types, transcription factors (TFs), or target genes (TGs); edges represent intercellular communications or cell type-specific regulatory mechanisms, **b,** Summary statistics about the number of edges (left) and nodes (right) of PFGs constructed by iBrainMap across 1,494 donors, **c,** Donors from multi-cohort AD studies: PsychAD (Mount Sinai Brain Bank/MSSM (M) (n=1,042), Rush Alzheimer’s Disease Center/RADC (R) (n=152), Seattle Alzheimer’s Disease Brain Cell Atlas/SEA-AD (S) (n=80) and NIMH-IRP Human Brain Collection Core/HBCC (H) (n=300). Data summary of phenotypic donors used in this study divided into three levels: Disease vs. Control (AD vs. Control, SCZ vs. Control, AD-DLBD vs. Control, AD-resilient vs. AD vs. Control), Disease Progression (Braak stages, CERAD, Cogdx), and Neuropsychiatric symptoms (Dysphoria, DecInt (Anhedonia), Sleep/Weight Gain/Guilt/Suicide (S/W/G/S), Depression/Mood (D/M)); each horizontal bar is a phenotype contrast.

**Extended Figure 2:**
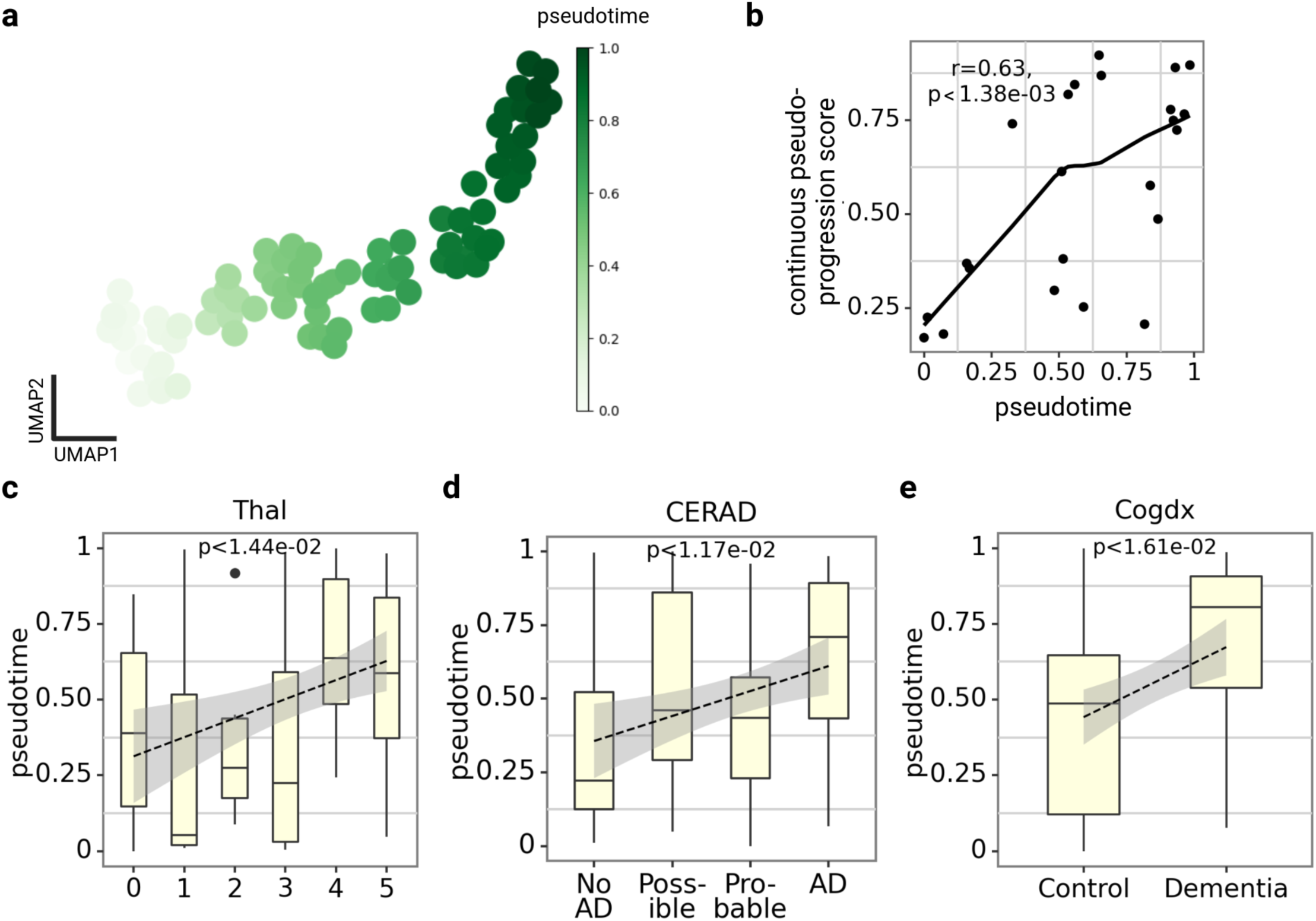
Independent validation of phenotypic pseudotimes by SEA-AD cohort. **a,** Graph embeddings are extracted for donors with different phenotypes using a pre-trained KG-GNN model to be used for pseudotime analysis and generate phenotypic pseudotimes, **b,** Comparison of pseudotime of graph embeddings from KG-GNN with CPS shows high Pearson correlation *r* = 0.63, *P <* 1.38e-3, **c-e,** Boxplots show increasing pseudotime (from a.) along AD pathology for Thal (Mann-Kendall, *P <* 1.44e-2) and CERAD (Mann-Kendall, *P <* 1.17e-2), and clinical diagnosis cogdx (Mann-Kendall, *P <* 1.61e-2).

**Extended Figure 3:**
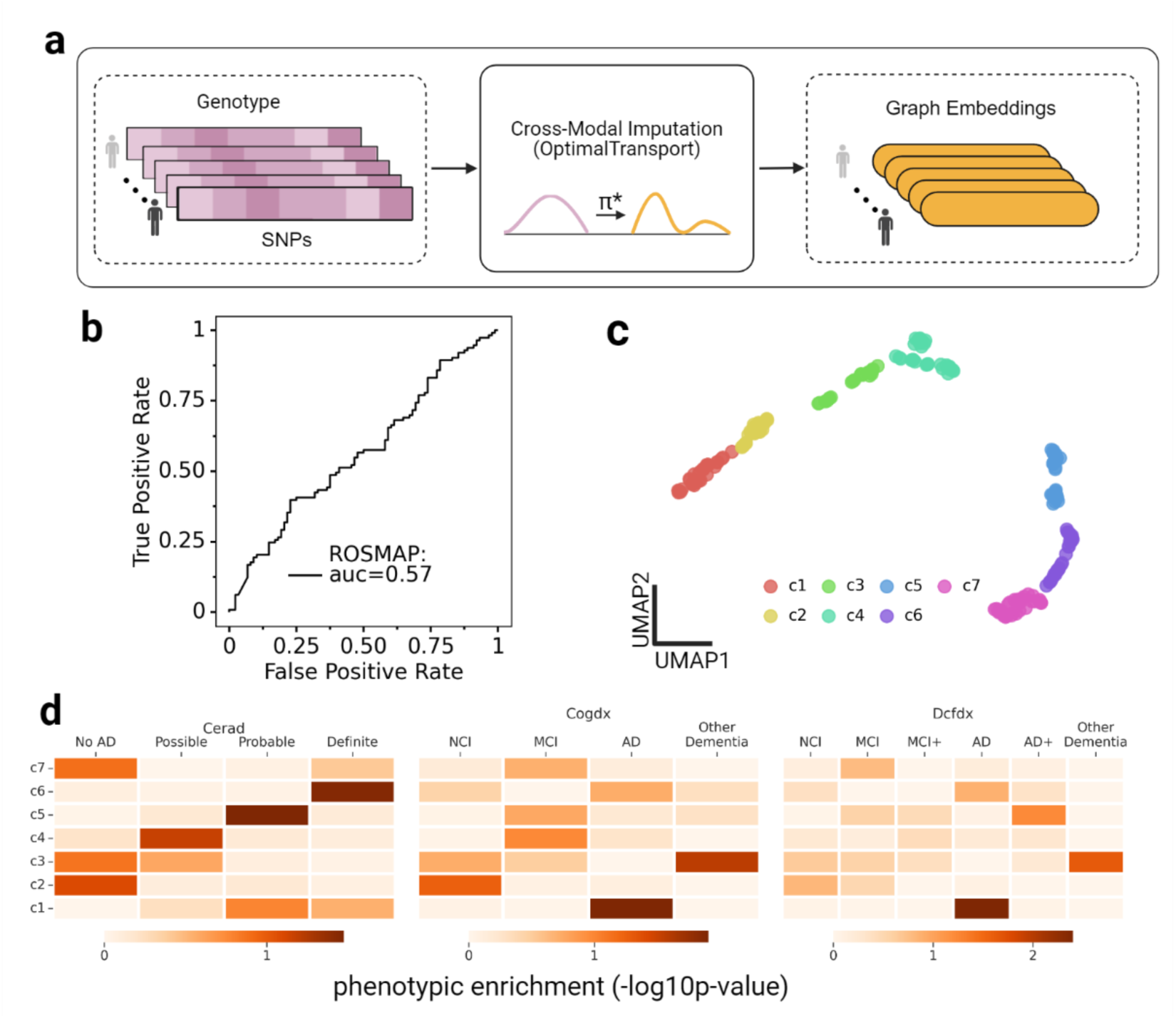
independent validation for graph embedding imputation from genotype. **a,** Cross-modal imputation of graph embeddings from genotype data using optimal transport^60^ (see **Supplementary Note 8**) **b,** ROC curve for classifying imputed graph embeddings into Early vs Late Braak stage donors from ROSMAP^59^ (Early (n=88) vs. Late (n=113) Braak stages) **c,** Graph embeddings based UMAP (Left: colored by Early vs Late Braak stage donors; Right: colored by clusters from unsupervised clustering/subtyping of graph embeddings) **d,** Heatmaps depict phenotype enrichments for clusters from **(c)** across AD progressions: AD pathologies (CERAD) and cognitive diagnosis (Cogdx, Dcfdx) showing an increasing trend from c1-c7, using -log10(hypergeometric test p-value) (see **Methods**).

## Notes

### Competing Interest Statement

The authors have declared no competing interest.

### Funding Statement

This study was funded by the National Institutes of Health grants, R01AG067025 (to P.R. and D.W.), RF1MH128695 (to D.W.), R21NS127432 (to D.W.), R21NS128761 (to D.W.), U01MH116492 (to D.W.), U01MH116442 (to P.R.), R01MH110921 (to P.R.), R01MH109677 (to P.R.), P50HD105353 (to Waisman Center), National Science Foundation Career Award 2144475 (to D.W.), Simons Foundation Autism Research Initiative pilot grant 971316 (to D.W.), and the start-up funding for D.W. from the Office of the Vice Chancellor for Research and Graduate Education at the University of Wisconsin-Madison.

