## Supplementary Materials for "Personalized Single-cell Transcriptomics Reveals Molecular Diversity in Alzheimer’s Disease"

### Table of contents

Supplementary Note 1: Graph Diffusion - p. 3

Supplementary Note 2: Knowledge-guided Graph Neural Network (KG-GNN) architecture - p. 3-4

Supplementary Note 3: Glossary of terms - p. 5-6

Supplementary Note 4: Training, Testing, and Validation - p. 6-7

Supplementary Note 5: Benchmarking machine learning algorithms for AD versus Control classification - p. 7-8

Supplementary Note 6: Classifying graph embeddings across AD phenotypes - p. 8

Supplementary Note 7: Node importance score computation - p. 8

Supplementary Note 8: Cross-modal imputation and classification of graph embeddings - p. 8

Supplementary Figures - p. 9-19

Supplementary Tables - p.20-22

References - 22

### Supplementary Notes

#### Supplementary Note 1: Graph Diffusion

We applied network diffusion to propagate well-known disease genes throughout the Personalized Functional Genomics graphs (PFGs) via its edges (see **Supplementary Figure 1**). Here we use the concept of insulated heat diffusion[1], where disease genes are treated as heat sources that retain some heat $\beta$ $(0 < \beta < 1)$ and equally distribute the rest to their neighboring nodes. The amount of heat propagated by disease genes, called diffused scores, represents the impact of these genes and are used as edge weights for the PFGs. The network diffusion requires two inputs: PFGs and prior knowledge (disease gene lists).

Let $G_i = (V_i, E_i)$ be the PFG for donor $i$, with $N_i = |V_i|$ nodes and $E_i$ representing the edges. Let $A_i \in \mathbb{R}^{N_i \times N_i}$ be the adjacency matrix and $D_i \in \mathbb{R}^{N_i \times N_i}$ is its diagonal matrix of the out-degree of nodes. The diffusion matrix $F_i$ is calculated as:

$$\mathrm{F_i} = \beta(\mathrm{I} - (1 - \beta)\,\mathrm{A_i D_i^{-1}})^{-1}, \quad (1)$$

where $I \in \mathbb{R}^{N_i \times N_i}$ is an identity matrix and $A_i D_i^{-1}$ represents the normalized adjacency matrix. Hence $F_i[t, s]$ represents the influence of a source node $s$ on target node $t$.

Given a collection of disease genes $d_g$, let $P_i \in \mathbb{R}^{N_i \times N_i}$ be a diagonal matrix encoding prior knowledge for an individual $i$:

$$P_i[j, j] = \begin{cases} 1, & \text{if gene } j \text{ is in } d_g \\ 0, & \text{otherwise} \end{cases} . \quad (2)$$

We combine the PFG and the prior knowledge to define the final edge weights for $G_i$ as

$$\mathrm{M_i} = F_i P_i. \quad (3)$$

We compute two versions of $M_i$: $M_{AD,i}, M_{SCZ,i}$ using our identified gene lists for AD and SCZ, respectively. We refer to $M_{AD}$ and $M_{SCZ}$ as *bio-diffused PFGs*, which are used to train our graph neural network model. Note that we can also compute these matrices for other gene-of-interest (GOI) lists ($M_{GOI}$). In this paper, we focus on AD and SCZ gene lists.

#### Supplementary Note 2: Knowledge-guided Graph Neural Network (KG-GNN) Architecture

KG-GNN uses a self-attention mechanism to calculate attention scores between a node and its neighbors (**Supplementary Figure 2**). These are normalized across the node's neighborhood using a softmax function and further used to update the node's features. This helps assign different weights to different neighbors, offering higher flexibility and interpretability to the model. For a Personalized Functional Genomics graph (PFG) $G_i$ (see **Supplementary Note 1**, **Supplementary Figure 1**), the set of node features $h^l = \{h_1^l, h_2^l, \ldots, h_{N_i}^l\}$, $h_t^l \in \mathbb{R}^{d_l}$, where $d_l$ is the dimension of node features at $l^{th}$ layer. The unnormalized attention coefficient $e_{t,s}^l$ between any node $t$ and its neighboring node $s$ is:

$$e_{t,s}^l = LeakyReLU\,(a^{l^T}\,(W^l h_t^l \,||\, W^l h_s^l)), \quad (4)$$

where $W^l \in \mathbb{R}^{d_{l+1} \times d_l}$ is a weight matrix, $a^l \in \mathbb{R}^{2d_{l+1}}$ is a learnable weight vector, and || denotes vector concatenation. We then compute the normalized attention coefficient $\alpha_{t,s}^l$ by normalizing $e_{t,s}^l$ across the node $t$'s neighborhood $\Omega_t$ using a softmax function:

$$\alpha_{t,s}^l = softmax\left(e_{t,s}^l\right) = \frac{exp(e_{t,s}^l)}{\sum_{n \in \Omega_t} exp(e_{t,n}^l)}, \tag{5}$$

where $\sum_{s \in \Omega_t} \alpha_{t,s}^l = 1$.

Before we update the node feature $h_t^l$, we incorporate prior biological knowledge through edges using our *bio-diffused PFGs* $M_{AD}, M_{SCZ}$ to get bio-diffused attention scores or priors $b_{(.),t,s}^l$:

$$b_{(.),t,s}^l = \frac{M_{(.)}[t,s]\alpha_{t,s}^l}{\sum_{n \in \Omega_t} M_{(.)}[t,n]\alpha_{t,n}^l}, \tag{6}$$

and specifically, the learned AD, SCZ and data-driven priors or bio-diffused attention scores are computed as follows:

$$b_{AD,t,s}^l = \frac{M_{AD,i}[t,s]\alpha_{t,s}^l}{\sum_{n \in \Omega_t} M_{AD,i}[t,n]\alpha_{t,n}^l}, \; b_{SCZ,t,s}^l = \frac{M_{SCZ,i}[t,s]\alpha_{t,s}^l}{\sum_{n \in \Omega_t} M_{SCZ,i}[t,n]\alpha_{t,n}^l}, \; b_{data-driven,t,s}^l = \frac{M_{data-driven,i}[t,s]\alpha_{t,s}^l}{\sum_{n \in \Omega_t} M_{data-driven,i}[t,n]\alpha_{t,n}^l},$$

where $\sum_{s \in \Omega_t} b_{(.),t,s}^l = 1$ and $M_{data-driven,i} = A_i \in \mathbb{R}^{N_i \times N_i}$ is the adjacency matrix of $G_i$.

Finally, the updated node feature for the node $t$ is computed by aggregating attention scores and passing it through a non-linearity $\sigma$ (sigmoid) over its neighbors:

$$h_t^{l+1} = \sigma\left(\sum_{s \in \Omega_t} b_{(.),t,s}^l W^l h_s^l\right). \tag{7}$$

For multi-head attention, the above operations are replicated $K$ times (each with different parameters), and the output is aggregated by adding feature-wise:

$$h_t^{l+1} = \sigma\left(\frac{1}{K}\sum_{k=1}^{K} \sum_{s \in \Omega_t} b_{(.),t,s}^{l,k} W^{l,k} h_s^l\right). \tag{8}$$

Here, we divided the number of heads $K$ into AD-driven, SCZ-driven, and data-driven: $K = k_{AD} + k_{SCZ} + k_{data-driven}$, and updated $h_t^{l+1}$ as follows:

$$h_t^{l+1} = \sigma\Big(\frac{1}{K}\Big(\sum_{k=1}^{k_{AD}} \sum_{s \in \Omega_t} b_{AD,t,s}^{l,k} W^{l,k} h_s^l + \sum_{k=1}^{k_{SCZ}} \sum_{s \in \Omega_t} b_{SCZ,t,s}^{l,k} W^{l,k} h_s^l$$

$$+ \sum_{k=1}^{k_{data-driven}} \sum_{s \in \Omega_t} b_{data-driven,t,s}^{l,k} W^{l,k} h_s^l\Big)\Big). \tag{9}$$

Once trained, the model outputs latent graph embeddings $z_i \in \mathbb{R}^{d^L}$, where $d^L$ is the dimension of the final $L^{th}$ layer, for each *bio-diffused PFG* $G_i$ for an individual $i$. This is computed by averaging all the node features in the $L^{th}$ layer of KG-GNN:

$$z_i = \frac{1}{N_i}\sum_{t=1}^{N_i} h_t^L. \tag{10}$$

The KG-GNN model optimizes the following binary cross-entropy loss across all $J$ samples:

$$L = \frac{1}{J}\sum_{i=1}^{J} (y_i log(MLP(z_i)) + (1 - y_i) log(1 - MLP(z_i))), \tag{11}$$

where we train a Multi-Layer Perceptron ($MLP$) to classify $z_i$ into AD versus Control class.

#### Supplementary Note 3: Glossary of Terms

In this work, iBrainMap performed analysis on multiple phenotype contrasts split into three main levels: Disease vs. Control, Disease Progression, and Neuropsychiatric Symptoms. This section provides their definitions.

**Disease vs. Control**

1. **AD vs. Control**

This contrast compares donors with Alzheimer's disease and their controls. Donors with AD are defined as those who have CERAD scores of 2, 3, or 4, Braak stage of 3 and above, and clinically proven dementia. Their respective control group is defined as one donor with a CERAD score of 1 and Braak stage within 0-3.

2. **SCZ vs. Control**

SCZ is any individual with SCZ diagnosis (*SCZ* | *Schizoaffective_bipolar* | *Schizoaffective_depressive*) and secondary diagnosis is not allowed, except for metabolic and eating disorders.

3. **AD-DLBD vs. Control**

DLBD is any individual with DLBD diagnosis (*DLBD*), and secondary diagnosis can be only AD.

4. **Pathology-cognition (AD-resilient vs. AD-strict vs. Control)**

This contrast integrates pathological and cognitive information to group donors into three categories: (1) Donors with a CERAD score of 4, Braak stages above 3, and clinically proven dementia (AD-strict), (2) Donors could have CERAD scores of 2, 3, or 4 and must not have dementia (AD-resilient), and (3) Donors with a CERAD score of 1 and Braak stage within 0-3 (Control).

**Disease Progression**

1. **Braak**

This contrast compares AD progression via Braak stages that measure neurofibrillary tangles, irrespective of donors' clinical diagnosis.

2. **Cognitive Dementia Rating Score**

This contrast compares the clinical dementia rating score (CDR score) where (0, 0.5, 1) = Control, (2, 3) = Mild Cognitive Impairment (MCI), and (4, 5) = Dementia. For SEA-AD[2], we renamed their Cognitive status phenotype as follows: No Dementia=Control, Dementia=Dementia.

3. **CERAD**

This contrast compares AD progression via qualitative variables from neuropathological scoring where 1=no AD, 2=possible AD, 3=probable AD, and 4=definite AD. For SEA-AD, we renamed their CERAD phenotypes as follows: absent=no AD, sparse=possible AD, moderate=probable AD, Frequent=definite AD.

**Neuropsychiatric Symptoms**

1. **Depression/Dysphoria vs. Control**

This contrast corresponds to depression and mood dysphoria. One donor is considered to be 'Case' - if only mood dysphoria appears to be true and 'Control' if mood dysphoria is not true and all other NPS corresponding to depression and mood are either NA or not true.

2. **DecInt vs. Control**

This contrast corresponds to depression and anhedonia. One donor is considered to be 'Case' - if only anhedonia appears to be true and 'Control' if anhedonia is not true and all other NPS corresponding to depression and mood are either NA or not true.

3. **Sleep/WeightGain/Guilt/Suicide vs. Control**

This contrast corresponds to sleep issues (early-, mid-, and late- insomnia, and hypersomnia), weight gain, guilt, and suicidal thoughts within AD lenient donors. One donor is considered to be 'Case' - if at least one of the above symptoms appears to be true and 'Control' if none of the symptoms are true.

4. **Depression/Mood vs. Control**

This contrast corresponds to depression and mood disorders. One donor is considered to be 'Case' - if at least one of the above symptoms appears to be true and 'Control' if none of the symptoms are true.

#### Supplementary Note 4: Training, Testing, and Validation

**Graph Subsampling:** We use the graph sampling technique, Neighbor Sampling[3], to sub-sample PFGs for training the KG-GNN model. In particular, we used the **`Neighborloader`** function from PyTorch Geometric[4] and set parameter **`num_neighbors`** to 10 neighbors to be sampled for each node for 100 iterations. This ensures connectivity of the subgraphs and information flow throughout the network. We set the **`batch_size`** (used for mini-batching) based on a hyperparameter to specify the number of subgraphs:

$$batch\ size = \frac{number\ of\ PFGs}{number\ of\ subgraphs}$$

We trained and tested our KG-GNN model on individuals from the MSSM cohort for binary classification of AD versus Control. Here stratified split individuals into 80% training and 20% held-out sets. To find the optimal hyperparameters, we performed a 5-fold cross-validation (CV) and evaluated our model's performance on the held-out set. We also tested our pre-trained model on an independent dataset from the RADC cohort.

**Cross-Validation:** For our model training and evaluation, we use a modified Cross-validation (CV) and testing scheme as the MD-AD model[5], in which we perform five separate rounds of model tuning with CV followed by evaluation in a test set. For a single round, one-fifth of all samples are assigned to a held-out test set. Then using the remaining 4/5ths of the samples, we perform a five-fold CV to select hyperparameters with the best prediction performance. We then train the selected model using the full training set (4/5ths of the original data) and then report performance on the held-out test set. In order to evaluate the robustness of our evaluation metrics under different splits, we initially split the full dataset into five separate groups and repeated the above process five total times, where each one-fifth of the data acted as a held-out test set once. We note that across these iterations, different training sets selected different configurations of hyperparameters, and for each train/test round, we trained the full training set on the specific configuration selected by CV in that training set.

**Hyperparameter Tuning:** We tuned our model over a range of hyperparameters: optimizer ∈ [Adam, SGD, Adagrad], learning rate ∈ [1e-4, 5e-4, 1e-3, 5e-3], weight decay ∈ [0, 5e-3, 5e-4], dropout ∈ [0.2, 0.3, 0.4, 0.5, 0.6, 0.7, 0.8], batch size ∈ [5, 10, 15], diffusion parameter $\beta$ ∈ [0.1, 0.3, 0.5, 0.7, 0.9], number of subgraphs ∈ [2, 3, 5, 10] and number of attention heads ∈ [2, 4, 6, 8], number of GAT layers in KG-GNN model ∈ [1, 2, 3], GAT input layer dimension ∈ [2048, 1024], hidden GAT layer dimensions ∈ [2048, 1024, 512, 256, 128, 64], MLP hidden dimensions ∈ [128, 64]. We show our model's performance under various hyperparameters in **Supplementary Figure 4.**

**Performance metrics:** We evaluated our model's performance using metrics such as balanced accuracy (BACC) and the area under the receiver operating characteristic curve (AUROC) to address the imbalanced nature of our training data. The following terms are used to compute BACC and AUROC in **Figure 2**, **Supplementary Figures 3, 4, 5, 6**:

$$BACC = \frac{sensitivity\ +\ specificity}{2}$$
$$sensitivity = TPR = \frac{TP}{TP\ +\ FN}$$
$$specificity = \frac{TN}{FP\ +\ TN}$$
$$FPR = \frac{FP}{FP\ +\ TN}$$

where $TP$ is true positive, $TN$ is true negative, $FP$ is false positive, $FN$ is false negative, $TPR$ is true positive rate, and $FPR$ is false positive rate.

Our final model was picked based on the best average performance on the held-out test set based on the BACC and AUROC metrics for the AD classification task. **Supplementary Table 1** reports the final model layers, hyperparameters, and training details.

#### Supplementary Note 5: Benchmarking machine learning algorithms for AD versus Control classification

We benchmarked our knowledge-guided graph neural network (KG-GNN) model with other state-of-art graph learning algorithms like graph attention network (GAT) and graph convolution networks (GCN) with the personalized functional genomics graphs (PFGs) as inputs for the AD donors vs. Controls classification task (see **Supplementary Figure 2a**). Additionally, we also benchmarked other machine learning algorithms like Support Vector Machine (SVM), Logistic Regression (LR), and Multi-Layer Perceptron (MLP) to classify average cell-type gene expression for each donor into AD vs. Control groups (see **Supplementary Figure 2b**). We ran these models using default settings from the Python package scikit-learn[6]. The results demonstrate that our model outperformed others giving high classification scores across the different metrics.

#### Supplementary Note 6: Classifying graph embeddings across AD phenotypes

We classified the graph embeddings into different AD phenotypes using machine learning algorithms like Support Vector Machines (SVM), Logistic Regression (LR), and Multi-layer Perceptron (MLP) using the Python package Scikit-learn[6]. To do this, we performed 5-fold cross-validation where the dataset was stratified split into training and held-out for each fold using a 4:1 ratio. We evaluated the performance using metrics like AUROC and BACC and picked the best model based on average AUROC across five folds. The results are shown in **Figure 2b** across several phenotypes for binary and multi-class classification tasks for donors from the MSSM cohort (**Extended Figure 1c**): (1) Binary classification: includes phenotypes SCZ, AD-DLBD, Dysphoria, DecInt, S/WG/G/S, D/M; (2) Multi-class classification: includes phenotypes Braak (early vs mid vs late stages), CERAD (No AD vs. AD Possible vs. AD Probable vs. AD), Cogdx (Dementia vs. MCI vs. Controls).

#### Supplementary Note 7: Node importance score computation

We use edge importance scores to derive node importance scores. For a node $v_i$, let $N_{in}$ be the indegree of the node and $N_{out}$be the outdegree. We first calculate the indegree and outdegree importance scores of the node using the following formula:

$$I_{in} = ln(1 + \sum_{j} N_{in} b_{k,ji})$$

$$I_{out} = ln(1 + \sum_{j} N_{out} b_{k,ij})$$

where $b_{k,j,i}$ and $b_{k,i,j}$ are incoming and outgoing importance scores of an edge between nodes $v_i$ and $v_j$ respectively and k can be AD-driven, SCZ-driven, data-driven attention heads. Then the importance score of a node is computed using the formula:

$$importance\ score = \lambda * I_{in} + (1 - \lambda) * I_{out}$$

where $\lambda \in [0, 1]$ is a parameter to balance the indegree and outdegree importance scores. We empirically set $\lambda = 0.3$ based on the weighted average of the average indegree and outdegree of all nodes to give equal importance to both incoming and outgoing importance scores.

#### Supplementary Note 8: Cross-modal imputation and classification of graph embeddings

Here we used an optimal transport-based approach, CMOT[7], to impute graph embeddings for genotype data (e.g. ROSMAP dataset). Here, we trained the imputation model using the graph embeddings generated by the pre-trained KG-GNN model and genotype data for individuals from the MSSM cohort. We tried to impute embeddings for early versus late Braak stages in the ROSMAP data (**Extended Figure 3**). To classify the imputed graph embeddings, we applied the sklearn.svm.SVC() function from scikit-learn[5].

### Supplemental Figures

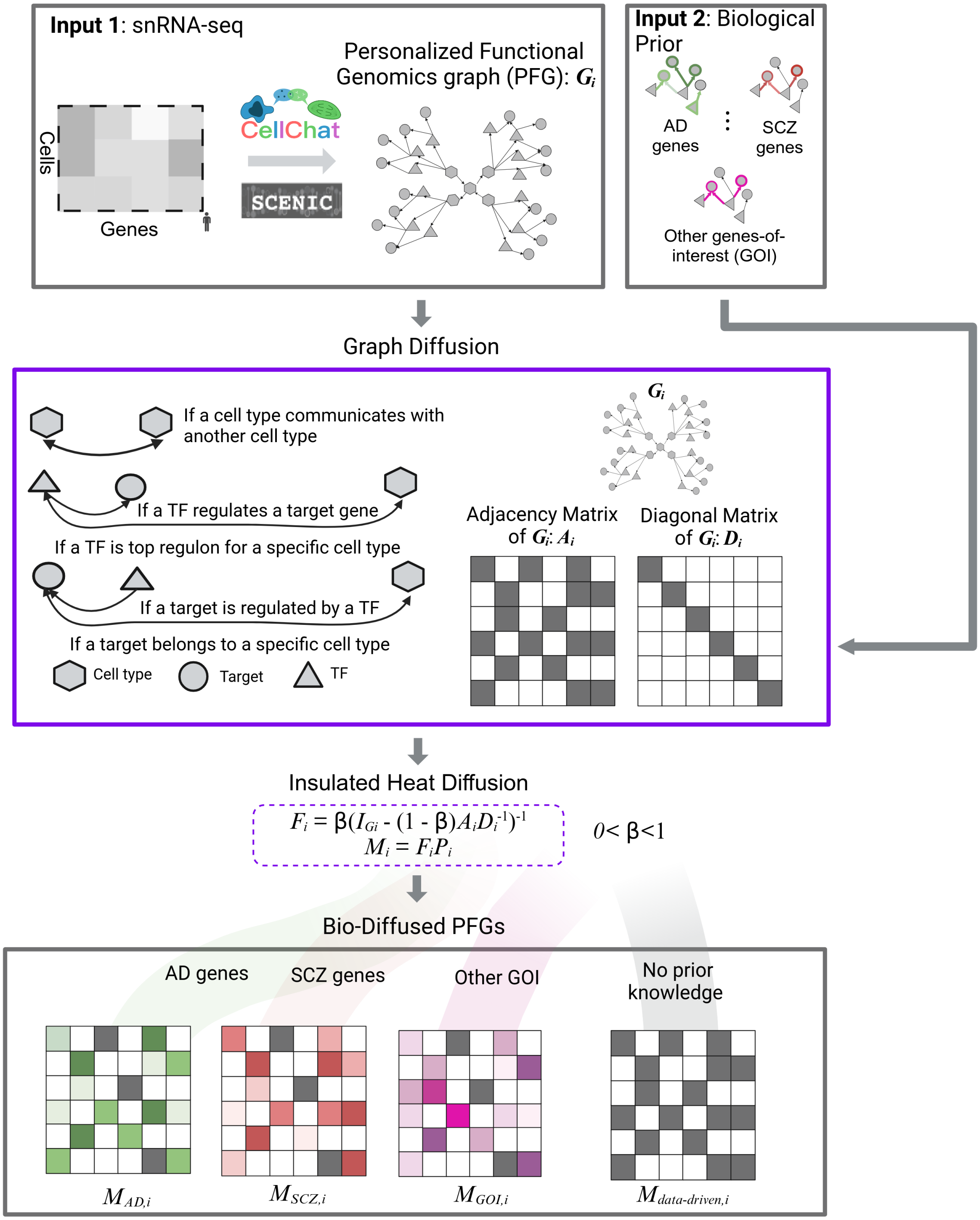


$$F_i = \beta(I_{Gi} - (1 - \beta)A_iD_i^{-1})^{-1}$$
$$M_i = F_iP_i$$

$M_{AD,i}$ $M_{SCZ,i}$ $M_{GOI,i}$ $M_{data\text{-}driven,i}$

**Supplementary Figure 1: Flowchart for constructing of bio-diffused PFGs.** *Bio-diffused* PFGs are constructed from two inputs: snRNA-seq of donors and biological (GOI)). First, personalized functional genomics graphs are built for each donor from their snRNA-seq using tools like CellChat and Scenic (see Supplementary Note 1). Each PFG has three node types: cell types, transcription factors (TFs), and target genes (TGs), connected by directed edges. Each edge captures distinct regulatory relations, for e.g., cell type interactions and cell type TF-TG regulation. We then computed *bio-diffused* PFGs using the concept of insulated heat diffusion for each donor with the help of adjacency and diagonal matrices of their PFGs. In particular, the resulting *bio-diffused* PFG $M_{AD,i}$, $M_{SCZ,i}$, and $M_{GOI,i}$ correspond to gene sets coming from known disease genes of AD, SCZ, and other genes-of-interests (GOI) for donor *i*. The $M_{data\text{-}driven,}$.is simply a matrix of ones.

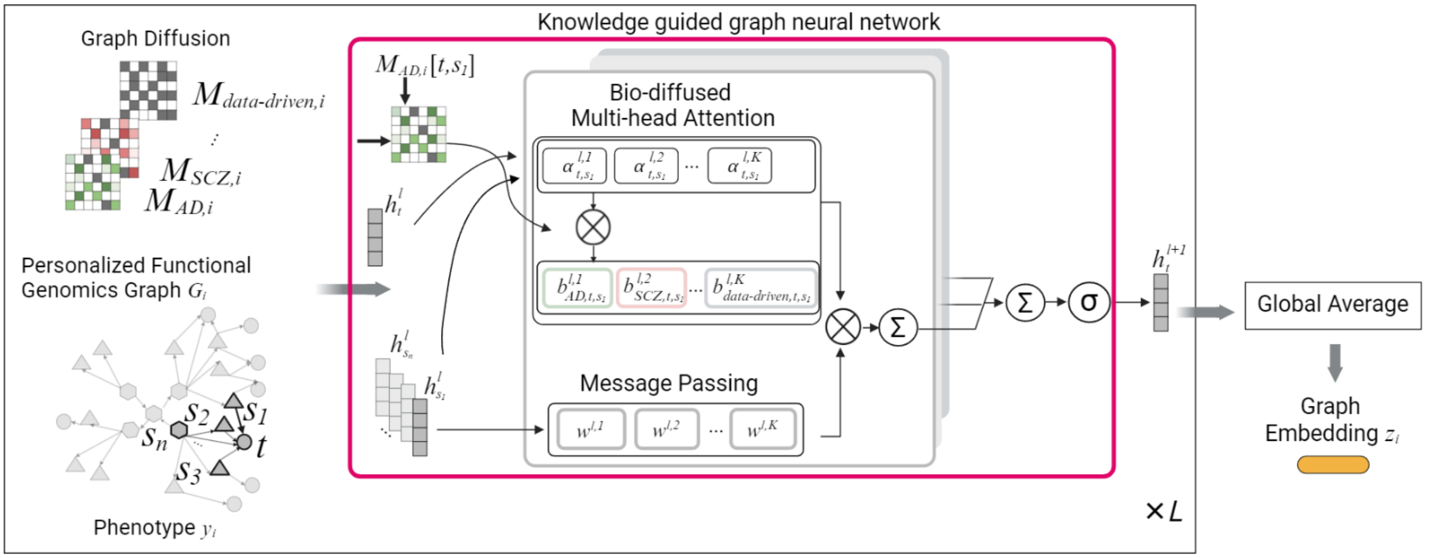


**Supplementary Figure 2: Architecture of knowledge-guided graph neural network (KG-GNN)** Given a PFG $G_i$, the inputs to the KG-GNN model include its *bio-diffused* PFG ($M_{AD,i}$, $M_{SCZ,i}$, ..., $M_{data\text{-}driven,i}$) and node features *h*. The KG-GNN uses a self-attention mechanism to calculate attention scores between a node *t* and its neighbors $s_j$ and incorporates prior biological knowledge through edges using our *bio-diffused PFGs* $M_{AD,i}$, $M_{SCZ,i}$ to get bio-diffused attention scores or priors $b_{AD,t,sj}$, $b_{SCZ,t,sj}$, …, $b_{data\text{-}driven,t,sj}$. These are normalized across node *t*'s neighborhood and are used to compute a linear combination of the node features corresponding to them using a softmax function to further update the *t*'s node features. (see **Supplementary Note 2**)

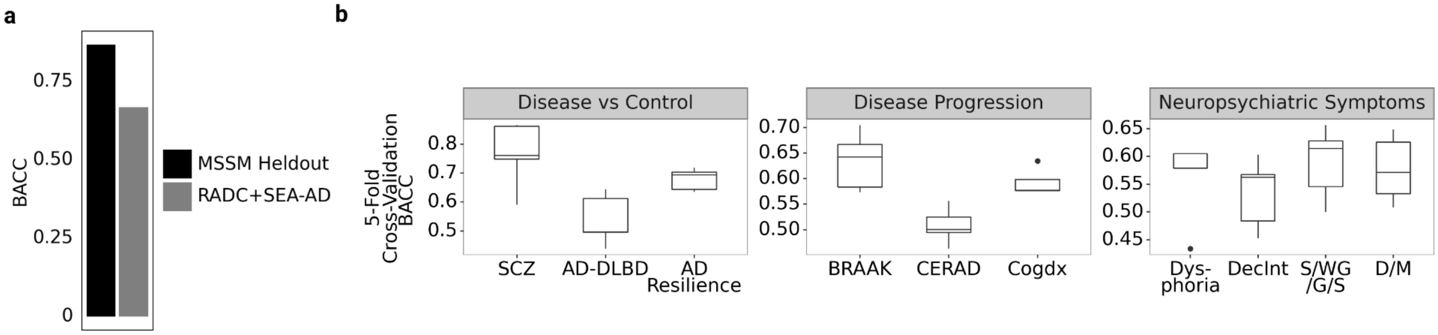


**Supplementary Figure 3: Graph embedding classifications for held-out data and disease pathology phenotypes. a**, Balanced accuracy (BACC) plot for classifying KG-GNN graph embeddings for AD donors vs. Controls in the MSSM held-out (AD (n=62) vs. Controls (n=30)) and RADC+SEA-AD (AD (n=93) vs Controls (n=68)) datasets described in Extended Figure 1c. **b**, Average five-fold cross-validated BACC for classification of KG-GNN graph embeddings across phenotype contrasts from **Extended Figure 1c**.

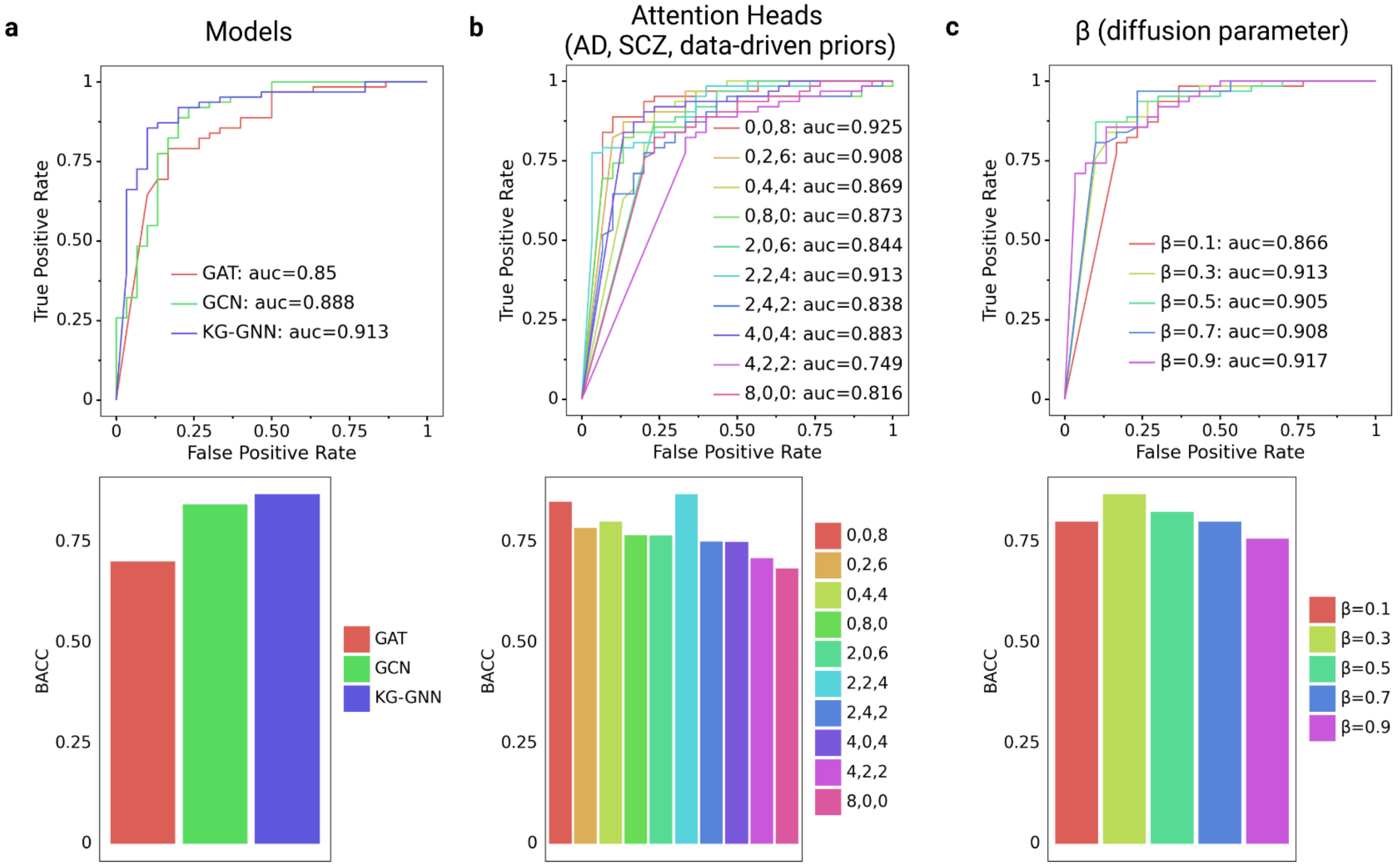


**Supplementary Figure 4: Benchmarking KG-GNN model performance for AD vs. Control classification a**, ROC curves and BACC comparing KG-GNN with state-of-art graph learning models: graph convolution networks (GCN), graph attention networks (GAT). **b**, ROC curves and BACC comparing different combinations of attention heads in the following order: AD, SCZ, Data-Driven. **c**, ROC curves and BACC comparing values for diffusion parameter $\boldsymbol{\beta}$. (see **Supplementary Note 4**)

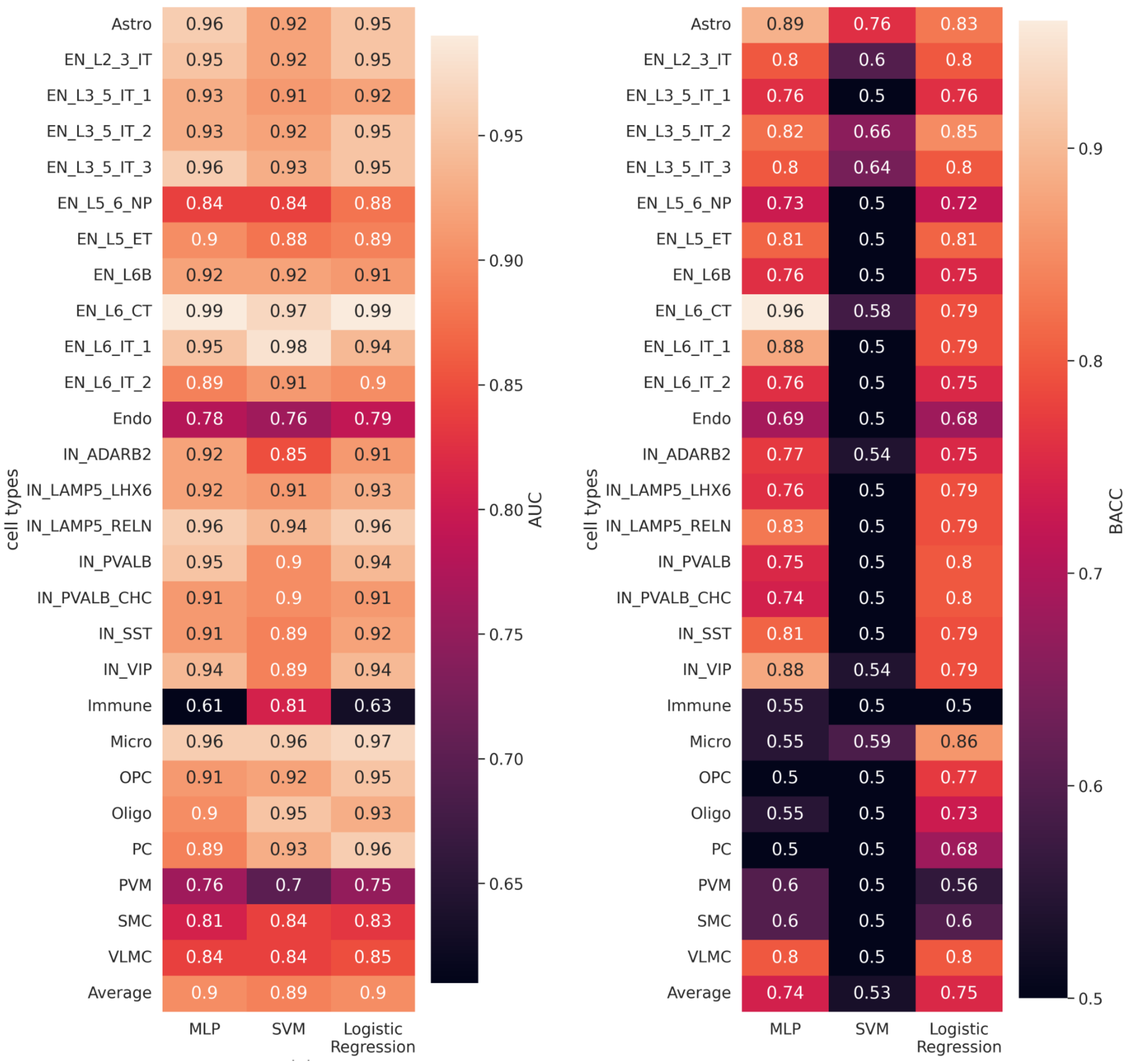


**Supplementary Figure 5: Benchmarking classification performance of donors with AD vs. Controls** for three machine learning models: Multi-linear Perceptron (MLP), Support Vector Machines (SVM), and Logistic Regression (LR). Here we classify each donor's average cell type gene expression into AD or Control groups using the three models across all cell types reporting metrics area-under-the-curve (AUC) and balanced accuracy (BACC) (see **Supplementary Note 5**).

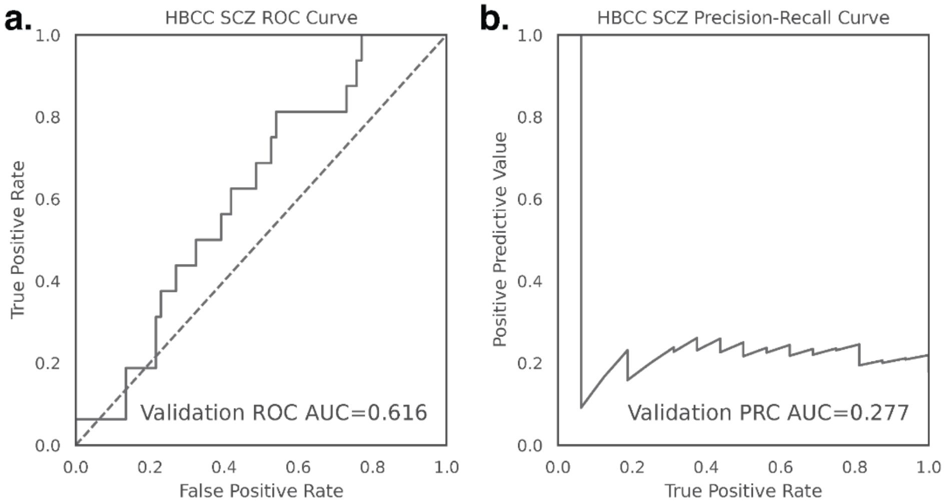


**Supplementary Figure 6: Predicting SCZ on HBCC using graph embeddings from the iBrainMap model. a,** ROC curve of predicted SCZ classification. **b,** Precision-recall curve of predicted SCZ classification.

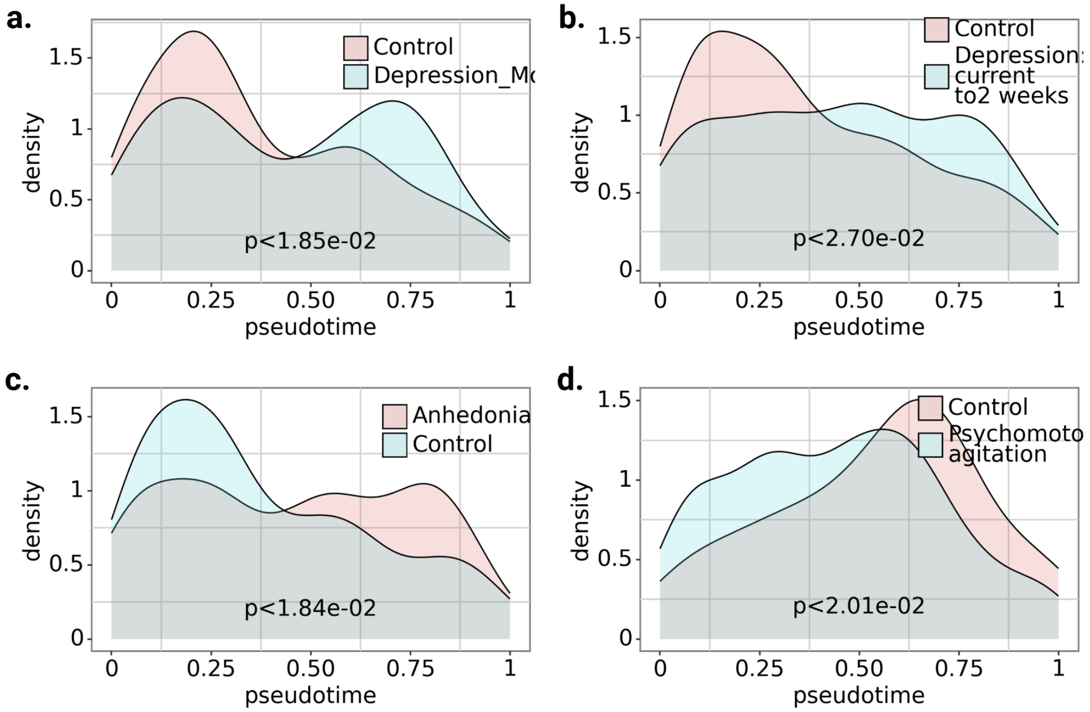


**Supplementary Figure 7: Density plots showing the distribution of neuropsychiatric symptoms (NPS) across individuals a,** Depression: Mood (P < 1.85e-02), **b,** Depression: current to 2 weeks (P < 2.70e-02), **c,** DecInt (Anhedonia) (P < 1.84e-02), and **d,** Psychomotor agitation (P < 2.01e-02) across NPS phenotypic pseudotimes.

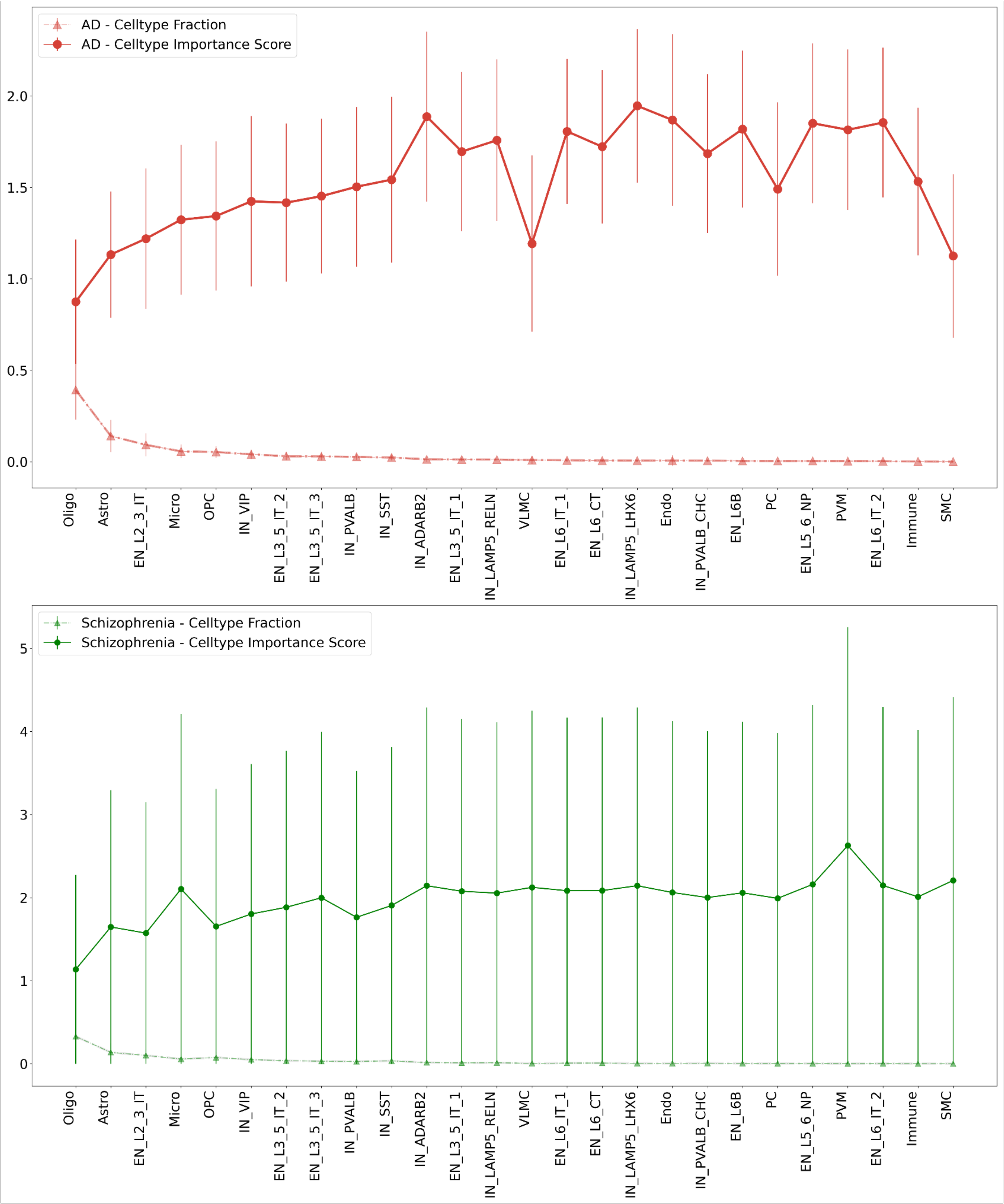


**Supplementary Figure 8: Cell type importance scores for AD (top) and SCZ (bottom) samples.** Circles indicate importance scores, and triangles represent cell fractions.

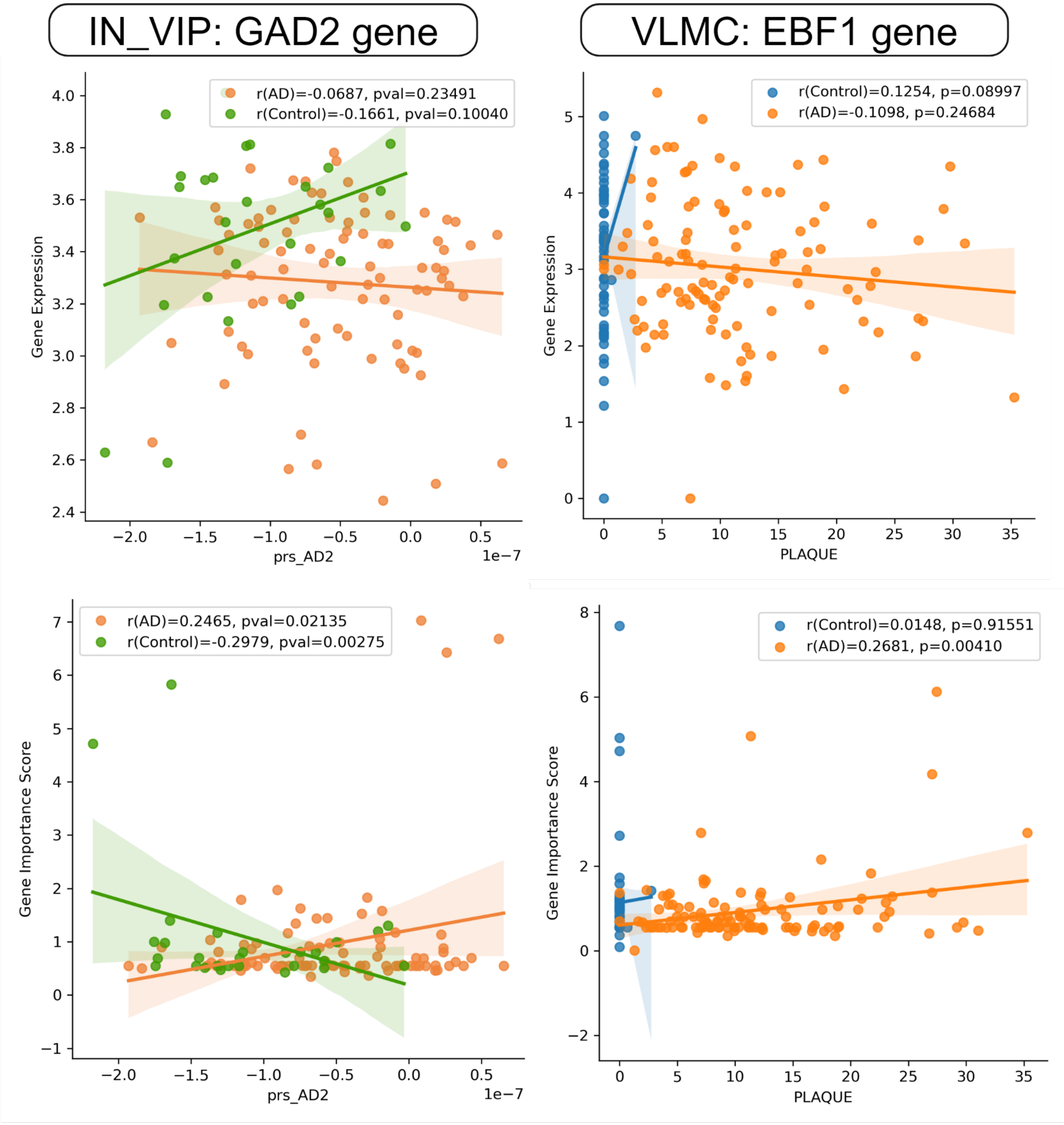


**Supplementary Figure 9: Correlation comparison of gene importance score and gene expression of some select genes.**

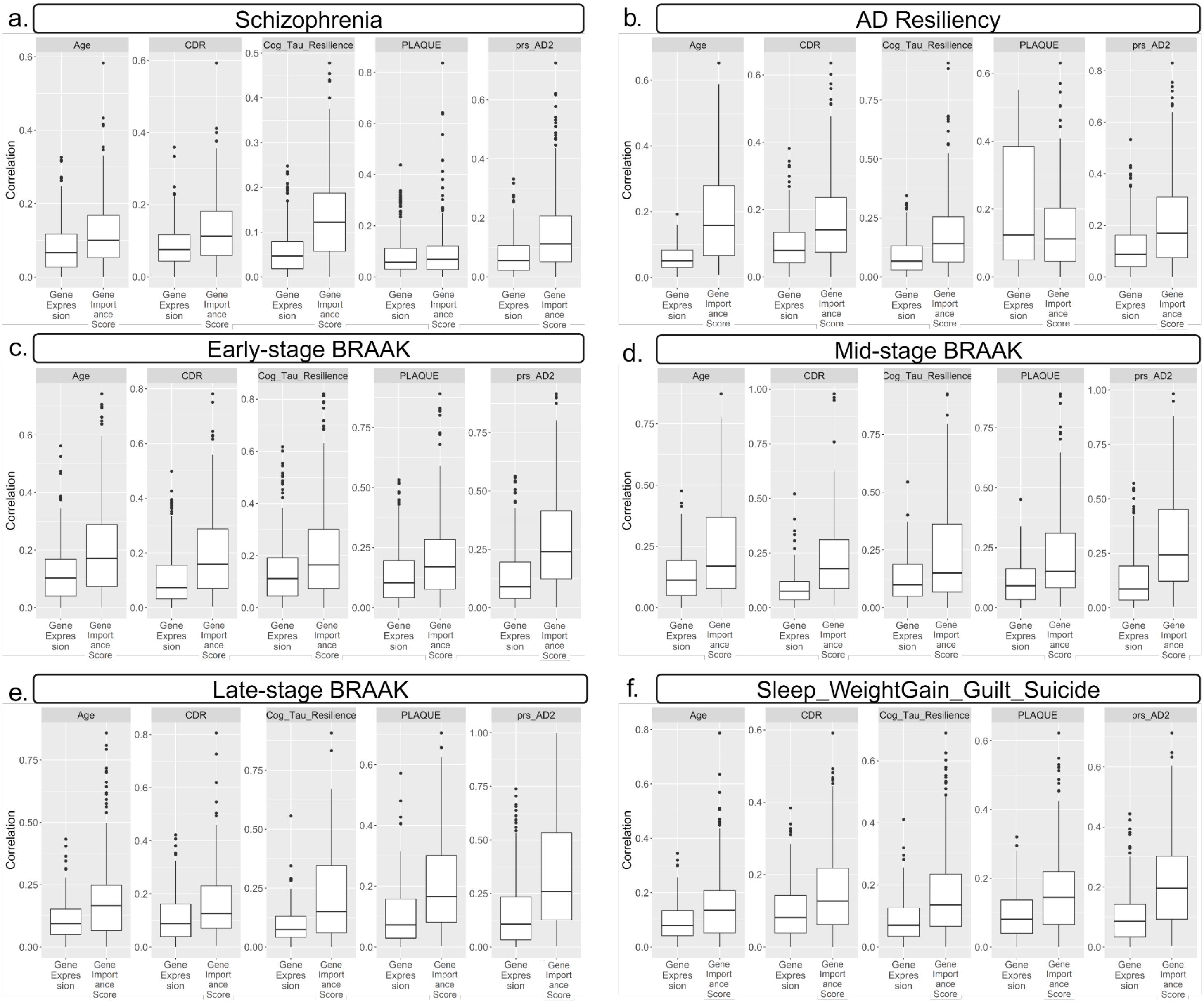


**Supplementary Figure 10: Correlation comparison of gene importance score and gene expression across multiple phenotypes.**

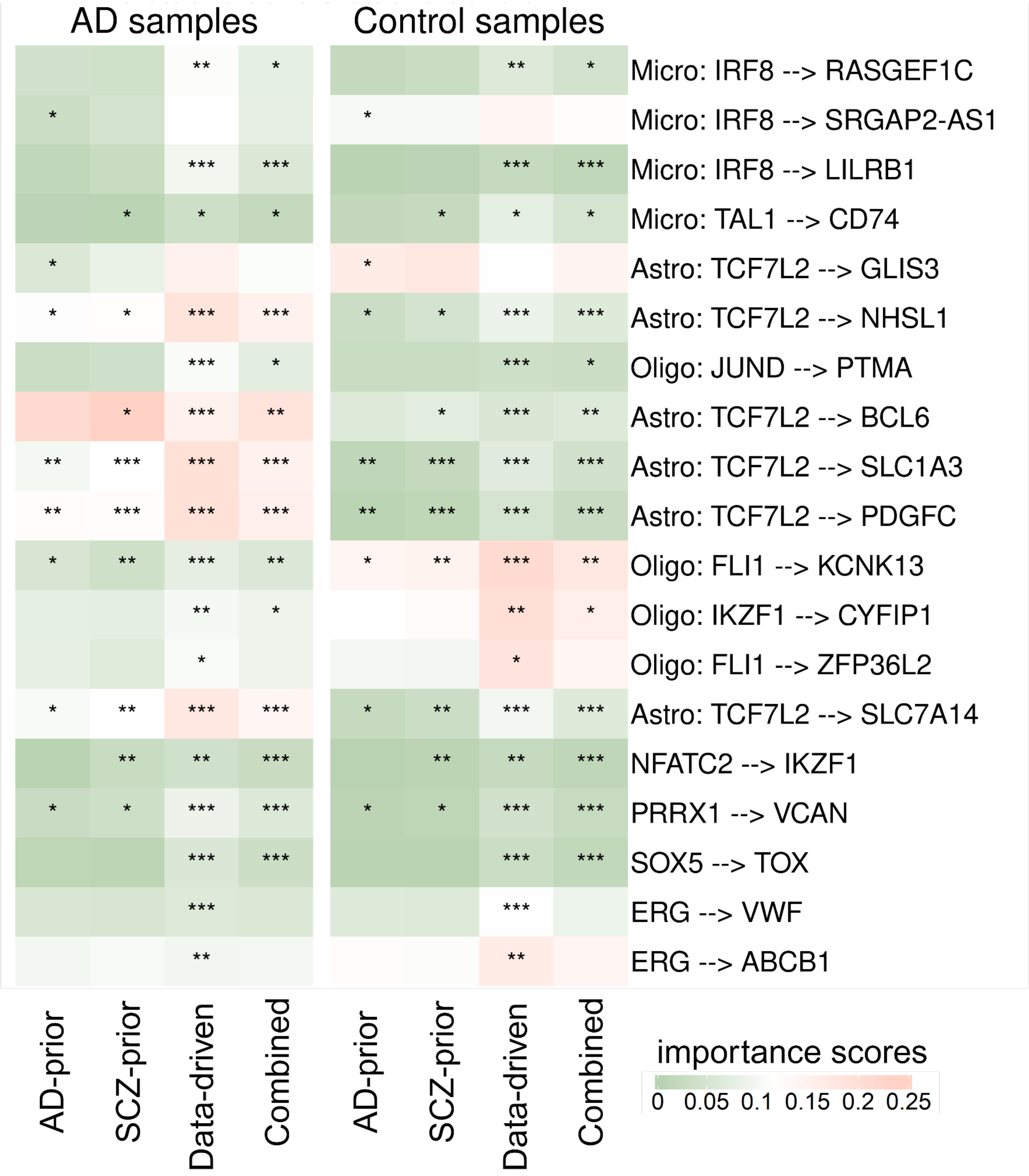


**Supplementary Figure 11: Importance scores for cell type TF-TG links between AD and control based on different priors.** Different significance levels are shown using the number of stars (* = p-value < 0.05, ** = p-value < 0.01, *** = p-value < 0.001).

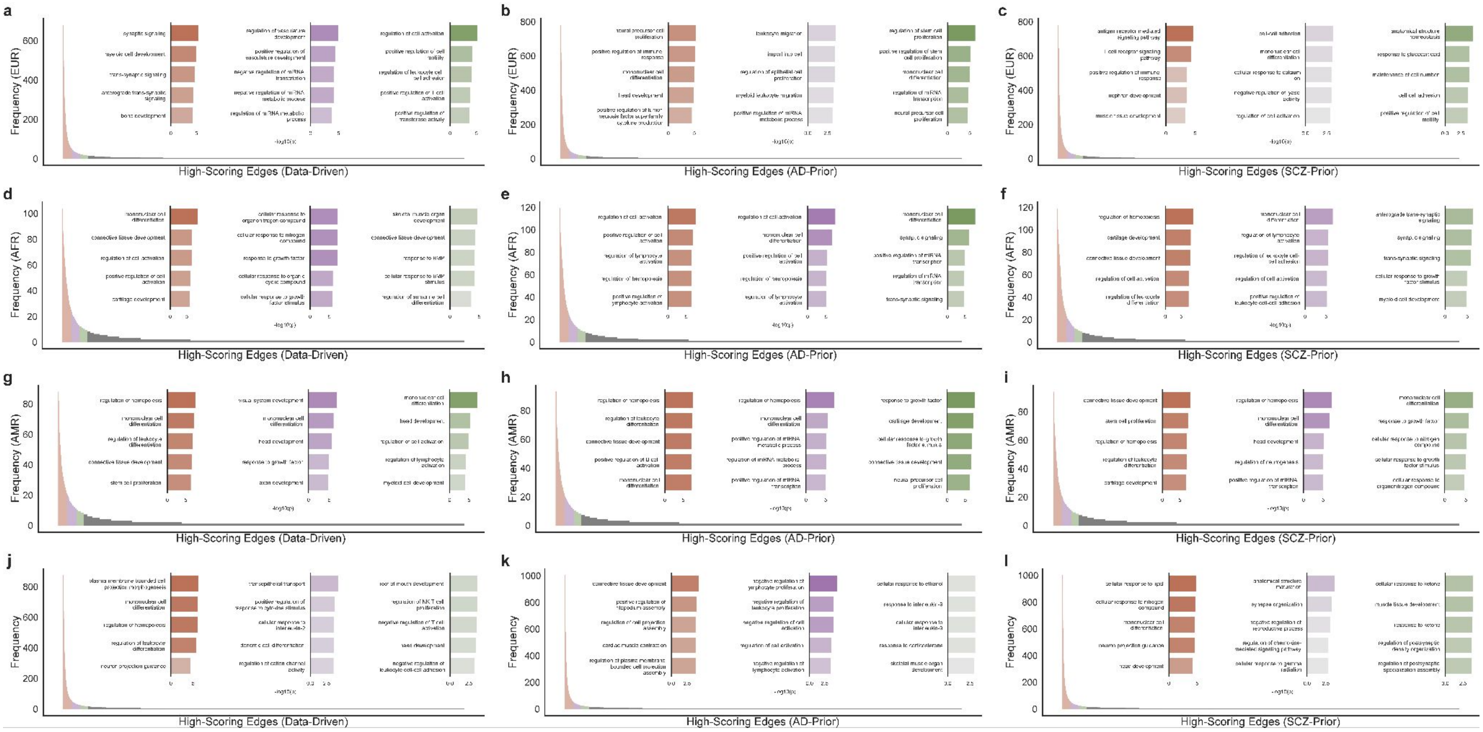


**Supplementary Figure 12: Determination of conserved and individual genes across ancestries. a-c,** Histogram of edges scoring in the top 5% of importance scores among the European (n = 792) population and GO-term enrichments (bar plots) for genes in the resultant highly conserved (top 2%, orange), conserved (top 2-4%, purple), and unique (top 4-6%, green) edge groups. **d-f,** The same population analysis in (a-c) repeated for African (n = 120) populations. **g-i,** The same population analysis in (a-c) repeated for admixed American (n = 99) populations. **j-l,** (a) repeated for the whole dataset. More details for this approach can be found in Supplementary Notes 4.2 and 4.3.

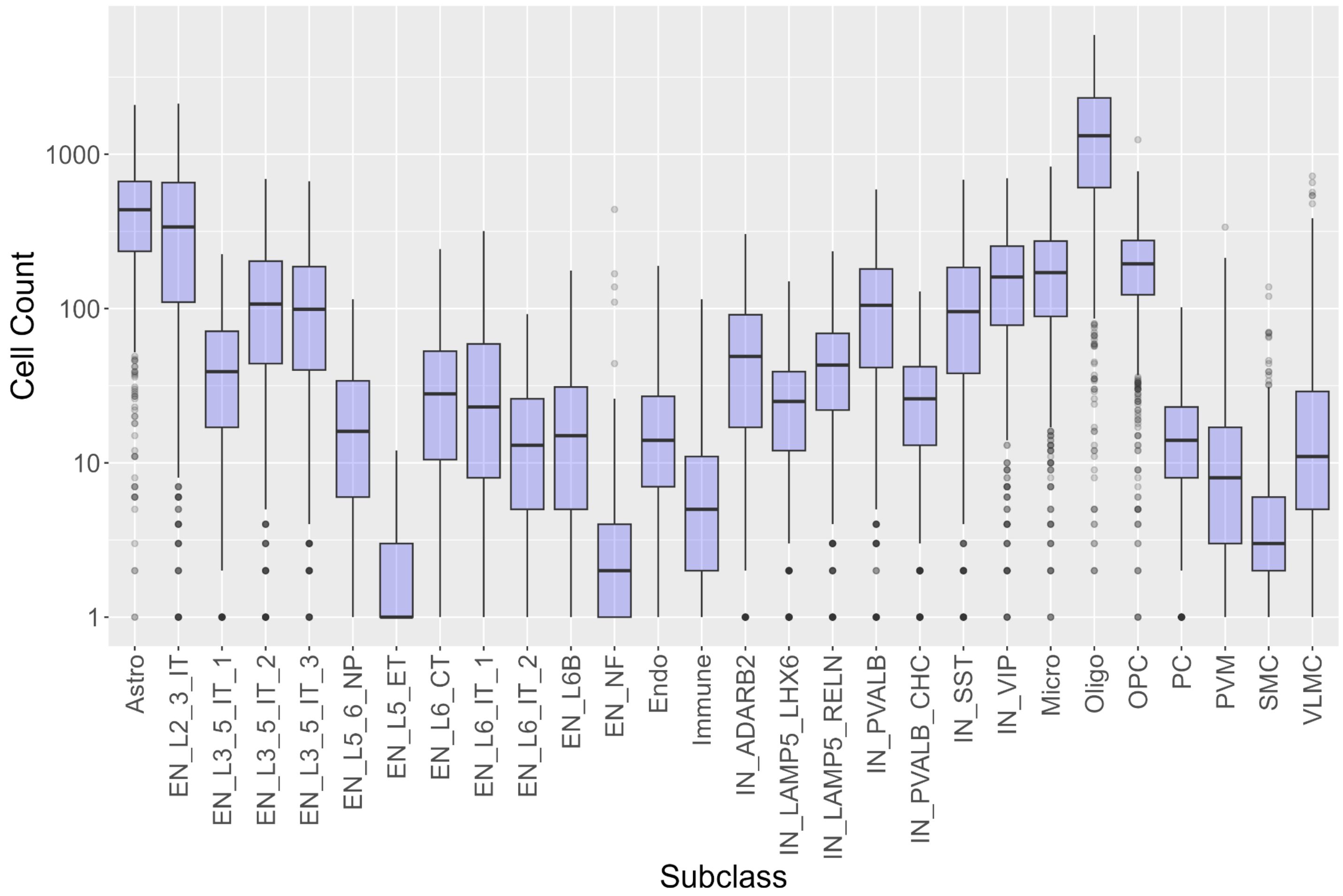


**Supplementary Figure 13: Cell counts per cell type across all donors.** The X-axis indicates the cell types, and the y-axis is the cell counts in the log scale for each cell type.

### Supplemental Tables

| **Hyperparameters:** | |
|---|---|
| Learning rate | 1e-4 |
| Batch size | 5 |
| Diffusion Parameter ($\beta$) | 0.3 |
| Dropout | 0.6 |
| Number of subgraphs | 3 |
| Number of attention heads | 8 |
| **Training:** | |
| Epochs | 23 |
| Memory (Mb) per epoch | 36,406 |
| Total runtime (s) | 1096.56 |
| **Model:** | |
| GATConv–>BatchNorm–>LayerNorm | 4 (2048, 1024, 512, 256) |
| Linear–>ReLU–>Dropout | 2 (128, 64) |
| Linear–>softmax | 1 |

**Supplementary Table 1: KG-GNN final hyperparameters, training, and model details**

| Phenotype | Clinical Phenotype | mean_corr_ importance_score | mean_corr_ gene_expression | p-value |
|---|---|---|---|---|
| AD_c15x | Age | 0.0978 | 0.0644 | 2.01E-07 |
| AD_c15x | CDR | 0.1144 | 0.0819 | 6.29E-08 |
| AD_c15x | Cog_Tau_Resilience | 0.1274 | 0.0550 | 1.85E-30 |
| AD_c15x | PLAQUE | 0.0773 | 0.0590 | 0.0165 |
| AD_c15x | prs_AD2 | 0.1237 | 0.0656 | 7.78E-12 |
| AD_resiliency | Age | 0.2008 | 0.0615 | 1.22E-07 |
| AD_resiliency | CDR | 0.1657 | 0.0923 | 1.25E-21 |
| AD_resiliency | Cog_Tau_Resilience | 0.1811 | 0.0893 | 7.53E-21 |
| AD_resiliency | PLAQUE | 0.1547 | 0.2037 | 0.0311 |
| AD_resiliency | prs_AD2 | 0.2095 | 0.1103 | 6.58E-31 |
| EarlyInsom | Age | 0.2071 | 0.1187 | 3.00E-12 |
| EarlyInsom | CDR | 0.1960 | 0.1044 | 6.57E-14 |
| EarlyInsom | Cog_Tau_Resilience | 0.2173 | 0.1473 | 1.57E-05 |
| EarlyInsom | PLAQUE | 0.2085 | 0.1360 | 1.29E-07 |
| EarlyInsom | prs_AD2 | 0.2831 | 0.1364 | 1.79E-20 |
| LateInsom | Age | 0.2107 | 0.1123 | 7.95E-08 |
| LateInsom | CDR | 0.1735 | 0.1111 | 3.09E-05 |
| LateInsom | Cog_Tau_Resilience | 0.2229 | 0.0988 | 5.76E-10 |
| LateInsom | PLAQUE | 0.2145 | 0.1093 | 3.03E-09 |
| LateInsom | prs_AD2 | 0.3454 | 0.1688 | 3.03E-10 |
| MSSM_SWGS | Age | 0.1490 | 0.0924 | 3.78E-13 |
| MSSM_SWGS | CDR | 0.1521 | 0.0998 | 1.26E-12 |
| MSSM_SWGS | Cog_Tau_Resilience | 0.1678 | 0.0856 | 9.56E-25 |
| MSSM_SWGS | PLAQUE | 0.1579 | 0.0936 | 1.91E-18 |
| MSSM_SWGS | prs_AD2 | 0.2139 | 0.0999 | 4.43E-31 |
| MidInsom | Age | 0.2440 | 0.1370 | 3.95E-07 |
| MidInsom | CDR | 0.2275 | 0.0963 | 7.95E-12 |
| MidInsom | Cog_Tau_Resilience | 0.2487 | 0.1254 | 5.23E-08 |
| MidInsom | PLAQUE | 0.2248 | 0.1127 | 2.30E-08 |
| MidInsom | prs_AD2 | 0.2955 | 0.1375 | 7.19E-15 |

| | | | | |
|---|---|---|---|---|
| SCZ_c07x | Age | 0.1221 | 0.0802 | 3.60E-08 |
| SCZ_c07x | CDR | 0.1266 | 0.0865 | 1.18E-14 |
| SCZ_c07x | Cog_Tau_Resilience | 0.1342 | 0.0586 | 3.34E-48 |
| SCZ_c07x | PLAQUE | 0.1020 | 0.0892 | 0.1738 |
| SCZ_c07x | prs_AD2 | 0.1443 | 0.0740 | 1.02E-19 |

**Supplementary Table 2: Correlation comparison of gene importance score vs gene expression and different phenotypes.**
